# Dissecting Clinical Features of COVID-19 in a Cohort of 21,312 Acute Care Patients

**DOI:** 10.1101/2023.11.27.23297171

**Authors:** Cole Maguire, Elie Soloveichik, Netta Blinchevsky, Jaimie Miller, Robert Morrison, Johanna Busch, W. Michael Brode, Dennis Wylie, Justin Rousseau, Esther Melamed

## Abstract

COVID-19 has resulted in over 645 million hospitalization and 7 million deaths globally. However, many questions still remain about clinical complications in COVID-19 and if these complications changed with different circulating SARS-CoV-2 strains.

We analyzed a 2.5-year retrospective cohort of 47,063 encounters for 21,312 acute care patients at five Central Texas hospitals and define distinct trajectory groups (TGs) with latent class mixed modeling, based on the World Health Organization COVID-19 Ordinal Scale. Using this TG framework, we evaluated the association of demographics, diagnoses, vitals, labs, imaging, consultations, and medications with COVID-19 severity and broad clinical outcomes.

Patients within 6 distinct TGs differed in manifestations of multi-organ disease and multiple clinical factors. The proportion of mild patients increased over time, particularly during Omicron waves. Age separated mild and fatal patients, though did not distinguish patients with severe versus critical disease. Male and Hispanic/Latino demographics were associated with more severe/critical TGs. More severe patients had a higher rate of neuropsychiatric diagnoses, consultations, and brain imaging, which did not change significantly in severe patients across SARS-CoV-2 variant waves. More severely affected patients also demonstrated an immunological signature of high neutrophils and immature granulocytes, and low lymphocytes and monocytes. Interestingly, low albumin was one of the best lab predictors of COVID-19 severity in association with higher malnutrition in severe/critical patients, raising concern of nutritional insufficiency influencing COVID-19 outcomes. Despite this, only a small fraction of severe/critical patients had nutritional labs checked (pre-albumin, thiamine, Vitamin D, B vitamins) or received targeted interventions to address nutritional deficiencies such as vitamin replacement.

Our findings underscore the significant link between COVID-19 severity, neuropsychiatric complications, and nutritional insufficiency as key risk factors of COVID-19 outcomes and raise the question of the need for more widespread early assessment of patients’ neurological, psychiatric, and nutritional status in acute care settings to help identify those at risk of severe disease outcomes.

## Introduction

The coronavirus disease 2019 (COVID-19) pandemic has caused an estimated 645 million hospitalizations and over 7 million deaths globally.^1^ Although COVID-19 mainly affects the respiratory system, it can also cause multi-system damage.^2^ While multiple studies have identified risk factors for COVID-19 respiratory severity,^3–5^ the rates of non-respiratory complications remain unclear. Furthermore, most COVID-19 studies have focused on hospitalized patients,^6–9^ leaving a knowledge gap in the clinical features of non-hospitalized patients evaluated in acute care facilities, such as the emergency department (ED).

Additionally, critical questions remain regarding patient outcomes in association with different circulating SARS-CoV-2 strains between 2019 and 2022.^10^ Notably, Omicron strains have been linked to less severe disease compared to earlier strains, attributed to either lower virulence and/or immunity from vaccinations and prior infections.^11^ However, it remains undetermined how specific clinical features have differed in patients with Omicron compared to earlier strains and whether there is a difference in clinical co-morbidities across different SARS-CoV-2 strains.

To address these questions, we examined a 2.5-year retrospective cohort of 21,312 COVID-19 patients at five ED/hospitals in Central Texas. We identified 6 distinct patient trajectory groups (TGs) and their unique clinical features. We also assessed how varying SARS-CoV-2 strains affected patient outcomes by TG. Our results expand upon clinical risk factors for COVID-19 severity in both hospitalized and ED-evaluated patients and provide insights into salient clinical features for COVID-19 patient outcomes.

## Materials and methods

### Study Approval and Data Collection

UT Austin Institutional Review Board (IRB ID: 2020-04-0117) and Ascension Seton (CR-20-066) approved the study. Electronic Health Record (EHR) data were extracted for 21,312 COVID-19 patients in Central Texas from the Ascension Seton Hospital Network clinical data warehouse with support and data sharing processes of the Dell Medical School Enterprise Data Intelligence Team.^12^ The Central Texas Seton Hospital network consisted of 1 academic medical center and 4 community hospitals spanning Travis, Hays and Williamson counties. Patients were included based on having a COVID-19 (U07.1) diagnosis code with all events being referenced to the patient’s first acute care event related to their first COVID-19 diagnosis code. Funding sources did not have a direct role in design, analysis, or approval of this manuscript.

### World Health Organization Ordinal Scale

Patients were retrospectively scored daily based on oxygen status, adapting the World Health Organization (WHO) COVID19 Ordinal Scale (Fig. 1), ^13^ classified as: 1 = asymptomatic, 2 = mild limitation in activity, 3-4 = hospitalized with mild/moderate disease (3 = room air/4 = nasal cannula or facemask oxygen), 5-7 = hospitalized with severe disease (5 = high-flow nasal cannula or noninvasive ventilation, 6 = intubation/mechanical ventilation, 7 = intubation/mechanical ventilation and organ failure), 8 = death. Scores of 1 and 2 were combined into 1.5 due to lack of outpatient data to quantify limitation of activity.

**Figure 1:**
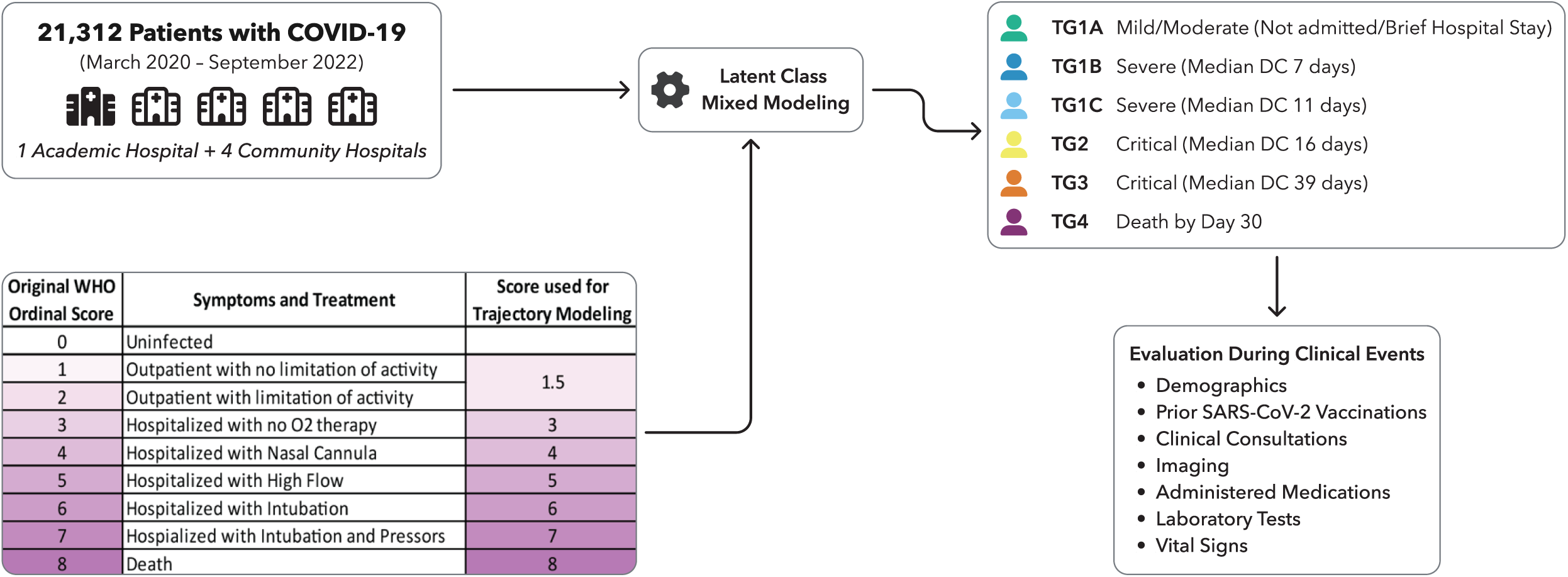
Experimental schema of analysis. Modeling of modified WHO ordinal scale identified six trajectory groups representative of increasing disease severity. Trajectory Group (TG) 1 consisted of mild/moderate patients who were evaluated in the ED, but not admitted; TG1B and TG1C represented severe hospitalized patients with short discharge; TG2 and TG3 were made up of critical patients with longer hospitalization, and TG4 was made up of fatal patients.

### Latent Class Mixed Modeling and Statistical Testing

Latent class mixed modeling (LCM) (R package lcmm (v2.0.0)) (Supplemental Fig. 1)^14^ was used to identify patient TGs for the modified WHO ordinal score over the course of 30 days, with additional details in the Supp. Methods.^3^ To differentiate TG1 trajectories, LCM was repeated with TG1 patients over a 12-day period, identifying TG1A/TG1B/TG1C subclasses. Central Texas reported COVID-19 case counts and vaccination data was retrieved from the county data portal.^15^ Waves 1-5 were divided by county-reported cases (Fig. 3A).

Generalized linear modeling was used to simultaneously evaluate all demographic variables reported in Table 1 while controlling for their interaction with age. Diagnoses, imaging, consultations, administered medications, laboratory tests and vital signs within the first two days of admission were evaluated via cumulative linked modeling using the ordinal R package (v2022.11.16).^16^ Longitudinal changes were evaluated with Generalized Additive Mixed Modeling using R package gamm4 (v0.2.6) (Table S10 and S11).^17^ Analyses controlled for hospital site, year/yearly quarter, sex, age, ethnicity, and race. For comparison of pairwise TG clinical features, significance was calculated using Chi-square testing. All analyses were Bonferroni adjusted for multiple hypothesis testing. Computational analyses were performed using the Biomedical Research Computing Facility at UT Austin, Center for Biomedical Research Support.RRID: SCR_021979.

**Table 1.**
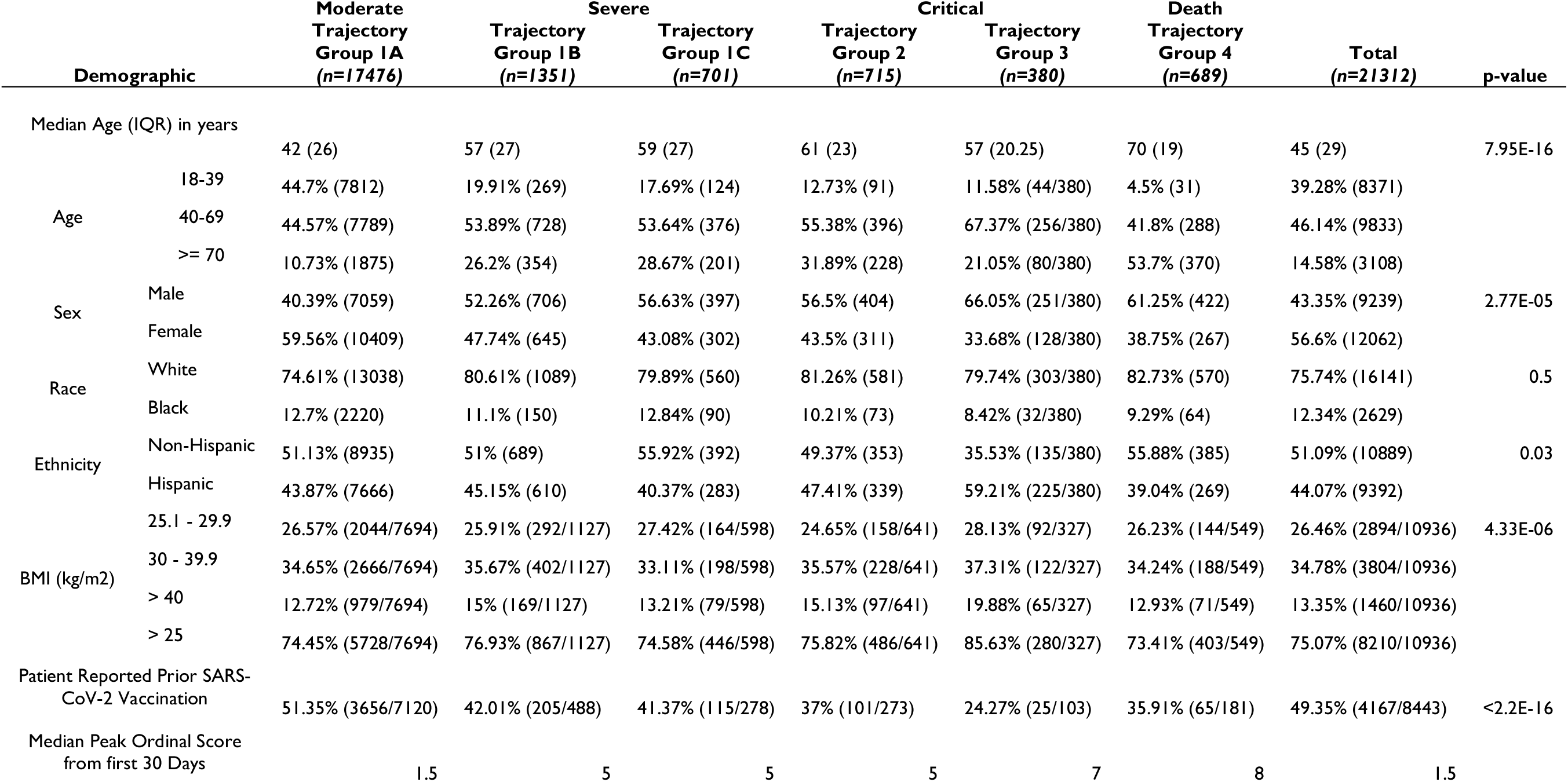
Demographics of Each Trajectory Group and Total Cohort.

### Data availability

Deidentified data will be made available upon reasonable request.

## Results

### Study Cohort

We analyzed 21,312 patient records across 47,063 retrospective encounters from five hospitals in Central Texas between March 2020 and September 2022 with the demographic features of the cohort displayed in Table 1 and Fig. 2. The 30-day mortality rate for all evaluated patients was 3.23%, with a mortality rate of 7.73% among only admitted patients, and a 17.96% mortality rate among only severe/critical patients (TG1B-TG4).

**Figure 2:**
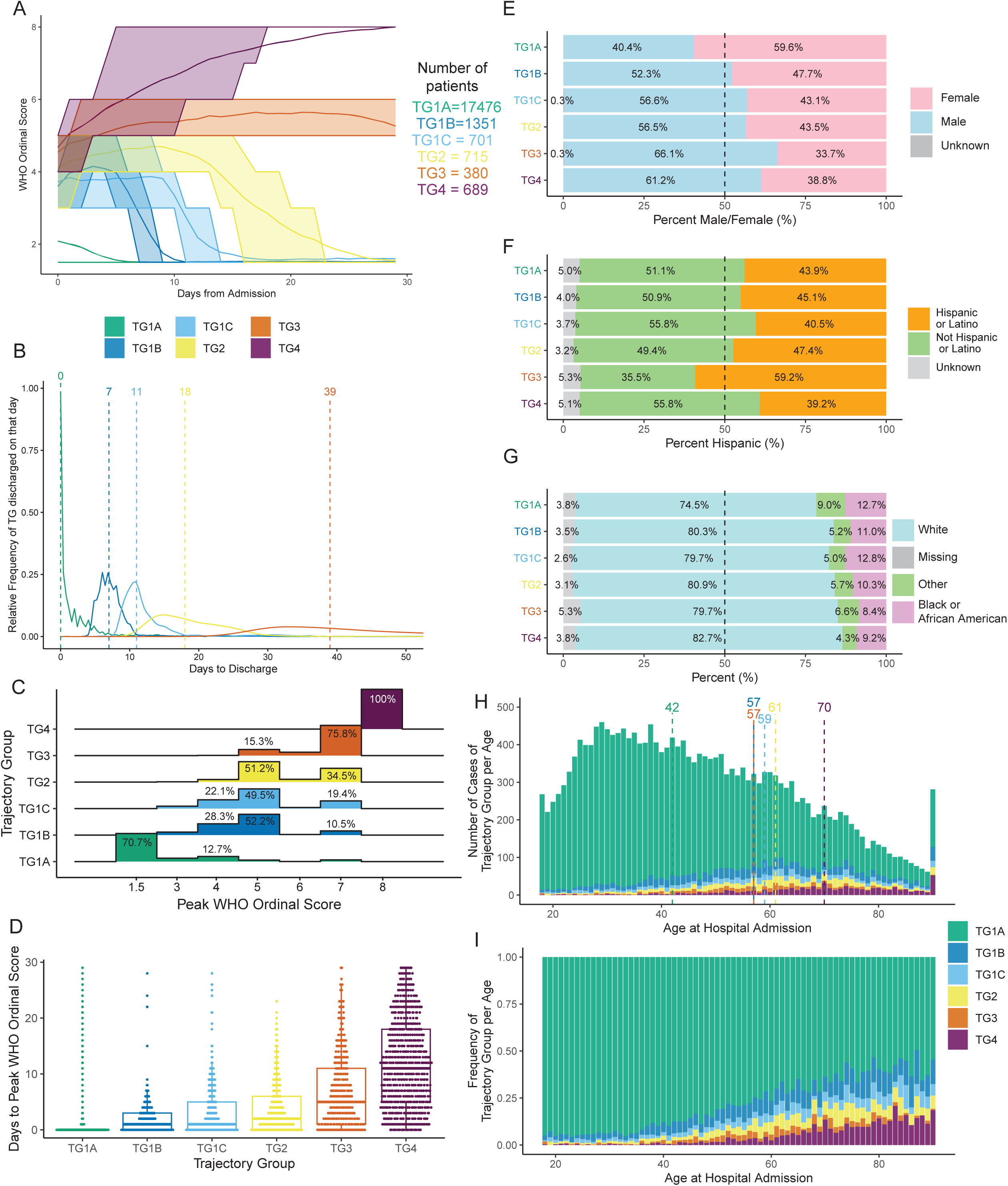
COVID-19 Trajectory Groups Capture Disease Severity and Reveal Underlying Demographic Differences. **(A)** The average trajectory of the WHO ordinal scale for each trajectory group over 30 days, with shading indicating the interquartile range. **(B)** Distribution by trajectory group of days to discharge with the median of each group indicated by dotted line. **(C)** Distribution of the peak scores from the first 30 days of all participants in each trajectory group. **(D)** Box and whisker plot of the days to each patient’s peak score by trajectory group. **(E)** Biological Sex, **(F)** Ethnicity, and **(G)** Racial composition of each trajectory group. **(H)** Histogram of the number of cases per trajectory group with age. Dotted lines denote the median age of each trajectory group. **(I)** Relative frequency of each trajectory group with age.

**Figure 3:**
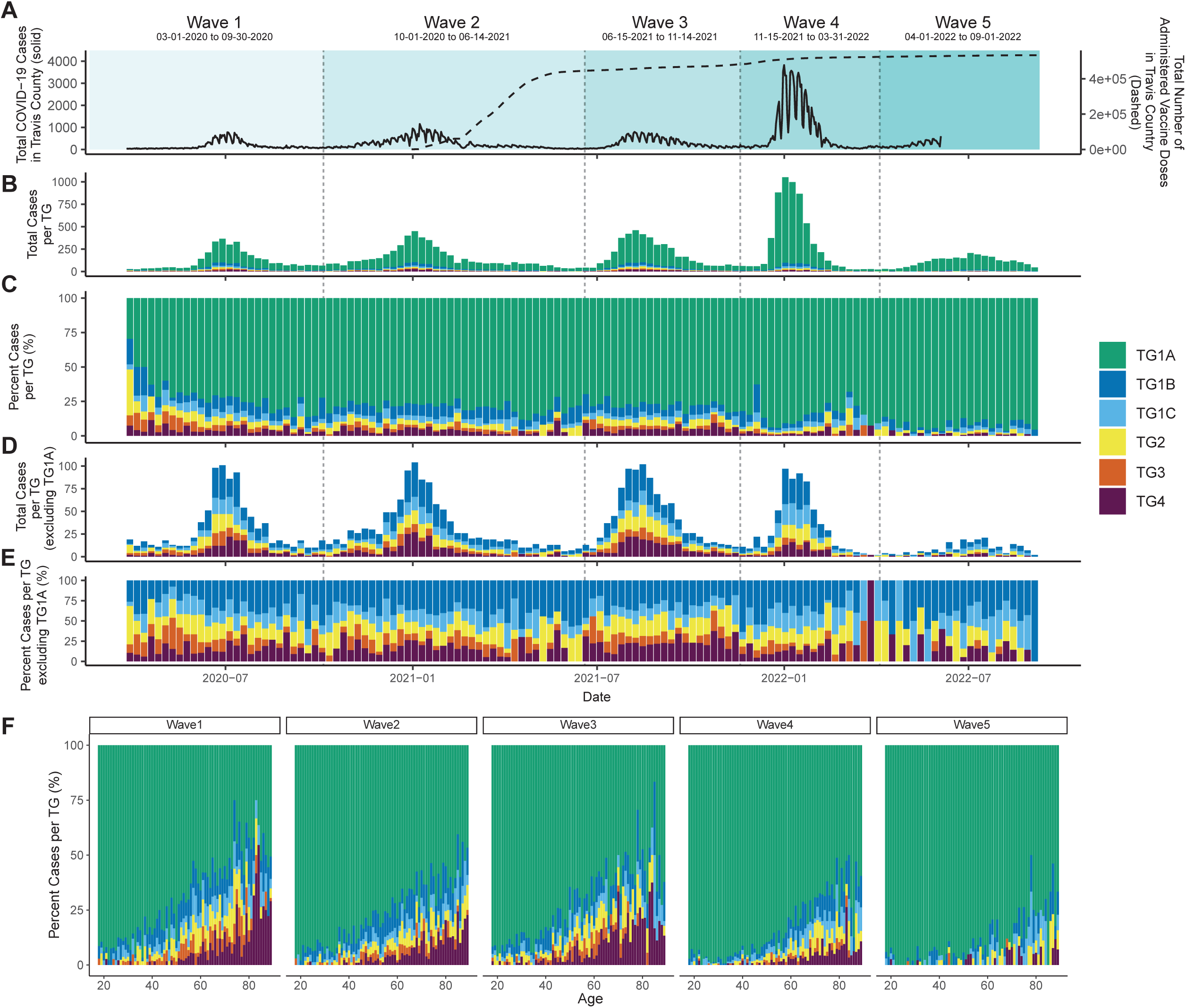
Trajectory Groups Over Time. **(A)** Total county reported COVID-19 cases (solid line, left axis) and vaccinations (dashed line, right axis) over the cohort sampling period. Waves 1-5 are denoted with their respective boundary dates. **(B)** Total cases per trajectory group by week across the sampling period. **(C)** Relative frequency of trajectory group for each week across the sampling period. **(D)** Total non-TG1A cases across each week and their **(E)** relative frequency. **(F)** Frequency of TG per year of age for Waves 1 through 5.

We identified four initial TGs on the modified WHO ordinal scale^13^ over 30-days (Fig. 2A). To better capture acute hospitalization trajectories, TG1 was further divided into TG1A, TG1B, and TG1C by repeating modeling over only 12 days (Supplemental Fig. 2). The TGs were reflective of patient time to discharge, peak ordinal score, and time to peak ordinal score (Fig. 2B-D). We used the following patient classification system - TG1A: patients with mild to moderate disease who were not admitted or were quickly discharged (median discharge of 0 days); TG1B: severe patients who required a moderate length of stay (median discharge of 7 days); TG1C: severe patients with a longer hospital stay (median discharge of 11 days); TG2: critical patients who discharged by day 30 (median discharge of 16 days); TG3: critical patients who were not discharged by day 30 (with a median discharge of 39 days); TG4: all patients died within 30 days. TG1 included 91.63% of all patients, with TG1A, TG1B, and TG1C comprising 82.00%, 6.34%, and 3.29%, respectively. TG2, TG3, and TG4 represented 3.35%, 1.78%, and 3.23% of the cohort.

TGs also displayed significant association with patient demographics including sex, ethnicity, race, and age (Fig. 2E&I, Table 1). Of note, males had worse outcomes, representing the majority of TG3 and TG4, while females had the highest representation in mild TG’s. Interestingly, patients documented as Latino/Hispanic were over-represented in TG3, however this difference was not observed in other TGs. There was no significant difference in race between TGs (Table 1, Fig. 2G). TG4 patients had the highest median age of 70, while patients in TG1A had the lowest median age of 42 (Fig. 2H). Notably, TG1B-TG3 had similar median ages ranging from 57-61.

When comparing patient-reported SARS-CoV-2 vaccinations, the mild TG1A had higher vaccinations compared to severe/critical TGs (Table 1). The lower severity groups TG1B and TG1C also had higher vaccination rates compared to critical groups TG2-TG4 (Table 1).

### Trajectory Groups Across COVID-19 Waves

Based on the total COVID-19 cases in Austin’s Travis County, we classified five different outbreaks or “waves” between March 2020 to September 2022 (Fig. 3A), roughly corresponding to Alpha/Beta (waves 1-2), Delta (wave 3), and Omicron (wave 4-5) (Fig. 3A), with the total number of hospitalizations reflective of the waves (Fig. 3B).

We observed significantly higher prevalence of TG1A in waves 3, 4, and 5 compared to waves 1 and 2 (Fig. 3C, Table S1). Additionally, despite higher total case counts during the Omicron spike (Wave 4), total severe and critical hospitalizations for TG1B-TG4 were comparable between all waves (Fig. 3D). Age-associated mortality also decreased across the waves (Fig. 3F).

In waves 4-5 (Omicron), we noted a reduction in pulmonary/critical care, palliative care, infectious disease, nephrology, and neurological consultations and a decrease in diagnoses of pneumonia, acute respiratory distress, type-2 diabetes, and hypertension, (Supplementary Table 2-3) supporting the trend of a higher proportion of mild patients (TG1A) in waves 4-5. Interestingly when evaluating only the severe/critical patients (TG1B-4), neurological and psychiatric consultations remained stable across all waves, suggesting that concern of neurological involvement in severe disease remained persistent across the different circulating SARS-CoV-2 strains (Fig. 4).

**Figure 4:**
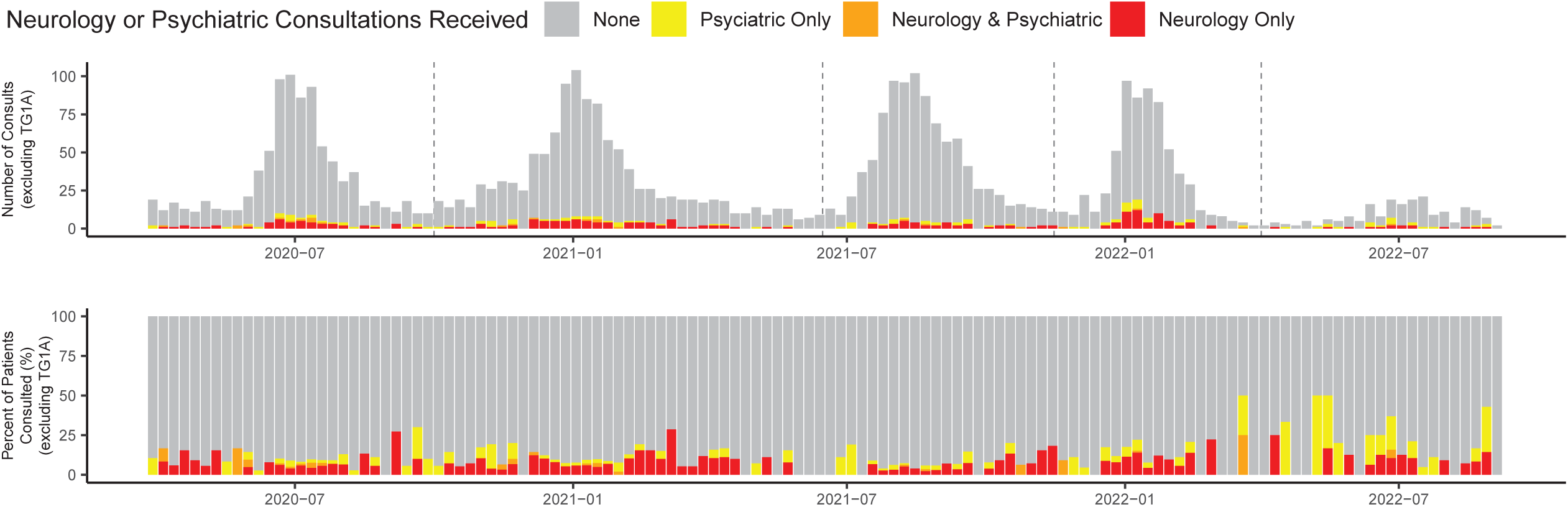
Neurological and Psychiatric Consultations Overtime. **(A)** Total and **(B)** percent of COVID-19 patients receiving Neurological and/or Psychiatric consultation(s).

### Diagnoses, Hospital Consultations, Imaging Studies, and Medications Across Trajectory Groups

#### Diagnoses

The comparison between the fatal (TG4) and most critical TG (TG3) revealed that TG4 patients had higher rates of cardiovascular co-morbidities, such as atherosclerotic heart disease, cardiac arrest, atrial fibrillation, nicotine dependence, and long-term aspirin use (Supplementary Table 4). Additionally, chronic kidney disease, cardiac arrest and chronic obstructive pulmonary disease were also elevated, implicating multi-organ damage in fatal patients compared to critical patients. Compared to TG4, TG3 patients were more likely to be anemic and experienced more electrolyte abnormalities and mixed acid base disorders. TG3 were also more likely to have ventilator dependence, sepsis, unspecified anxiety disorder, and dysphagia.

For the less severe groups, compared to TG1B-TG1C, TG1A patients had the least co-morbid diagnoses (Supplementary Table 4). Interestingly, TG1A patients had the highest association with headache and cough. Notably, alcohol related disorders were significantly higher in TG1B and TG1C patients compared to TG1A. Further, rheumatoid arthritis (RA) and inflammatory bowel disease (IBD), were significantly associated with TG1B-TG1C compared to TG1A. Additionally, severe/critical groups (TG1B-TG4) collectively had higher rates of several neurological and psychiatric disorders, including Alzheimer’s disease, cerebral infarction, epilepsy, migraines, myoclonus, neurocognitive disorder with Lewy bodies, non-traumatic intracerebral hemorrhage, Parkinson’s disease, restless leg syndrome, adjustment disorders, substance use related disorders, delirium, delusional disorders, depressive episode, generalized anxiety disorder, panic disorder, post-traumatic stress disorder, schizoaffective disorder, and schizophrenia (Fig. 5A). However, no neurological or psychiatric disorders were significantly different between the most critical TGs (TG3 vs TG2 or TG4 vs TG3).

**Figure 5:**
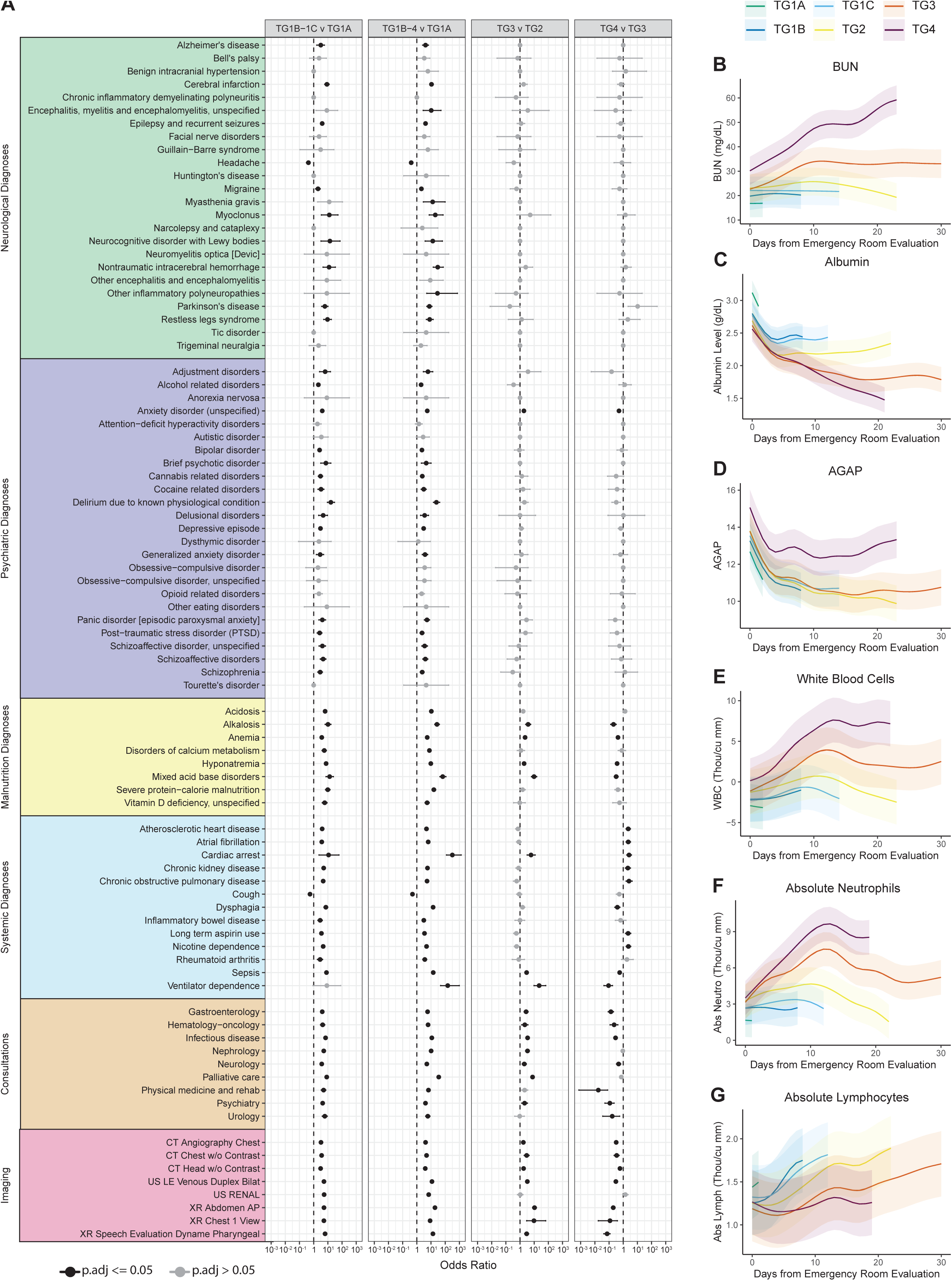
Diagnosis, Consultation, and Imaging Differences between COVID-19 TGs. **(A)** Odds ratios of key diagnoses, consultations, and imaging between TGs. Longitudinal modeling of laboratory testing over 30 days of hospitalization by TG for **(B)** Blood Urea Nitrogen (BUN), **(C)** Albumin, **(D)** Anion Gap (AGAP), **(E)** White Blood Cells, **(F)** Absolute Neutrophils, **(G)** Absolute Lymphocytes with 95% Confidence interval denoted as shaded region. Laboratory testing modeling results by TG were trimmed to days with data for greater than 10% of the TG.

#### Consultations & Imaging Studies

Although all consultations increased with higher TGs, TG4 received less critical care, infectious disease, gastroenterology, neurology, physical medicine and rehabilitation, hematology-oncology, psychiatry, and urology consultations compared to TG3 (Supplementary Table 5). Notably, TG3 and TG4 patients had the highest rates of palliative care and nephrology consultations, although only 64.20% of TG4 received a palliative care consult compared to TG3 (71.76%).

Notably, TG3 had the most imaging studies (Supplementary Table 6). Overall, most patients received respiratory imaging with increasing orders across TGs. Abdominal X-rays were significantly increased in more severe groups TG1B/TG1C compared to TG1A and in TG3 versus TG4. Furthermore, lower extremity venous ultrasound was also more common with increasing TG severity. Interestingly, 10.7% of the entire cohort received a Head CT without contrast, with frequency increasing from TG1A (7.6%) to TG3 (38.7%) (Fig. 5, Supplemental Table 6).

#### Medication Administration

The least medication administration was in TG1A, likely due to fastest discharge (Supplementary Table 7). Notably, dexamethasone usage was not different between TG3/TG4, while Remdesivir was higher in TG3. Higher medication usage in TG4 compared to TG3 included haloperidol, morphine, lorazepam, scopolamine, glycopyrrolate, which are often administered with comfort-care measures, in line with higher palliative care consultations and ’do-not-resuscitate’ orders in TG4. Additionally, neuromuscular blockers, atracurium, rocuronium, and succinylcholine as well as fentanyl, etomidate, and propofol were used more often in TG3 than TG4, correlating with a higher proportion of mechanical ventilation.

#### Laboratory Tests and Vital Signs

Several laboratory tests demonstrated significant separation of TGs at admission and through hospitalization (Fig. 5B-G). Interestingly, albumin was one of the best predictors of disease severity, displaying the most ordinal relationship across TGs, both at admission and throughout hospitalization (Fig. 4C, Supplementary Table 8), with lower values in the more severe TGs, in correspondence with higher malnutrition diagnosis in the critical TG3. On admission (day 0-1), several other predictive labs distinguished the mild TG1A group from the more severe groups. TG1A had higher red blood cell count, hematocrit, and hemoglobin compared with more severe TGs (Supplementary Fig. 4). In addition, TG1A had elevated absolute basophils, eosinophils, lymphocytes, lower absolute neutrophils and immature granulocytes compared to more severe TGs and lower D-Dimer, sedimentation rate, lactate, C reactive protein and fibrinogen as well as higher calcium compared to the more severe TGs (Supplementary Table 8). Furthermore, in comparing the critical and fatal TGs, TG4 had elevated AST, total bilirubin, creatinine, BUN and anion gap (AGAP), and lower total protein and CO2. Finally, the fatal TG4 had higher lactate, AGAP and lower arterial pH compared to critical TG3.

Vitals signs from the first two days were averaged for each patient to identify changes across TG (Supplementary Table 9). Notably, systolic and diastolic blood pressure and oxygen saturation decreased with severity along with higher respiratory rate and lower Glasgow coma scores (GCS). However only GCS was significantly different between TG4 and TG3. Over hospitalization, TG3/TG4 continued to have abnormalities in oxygen saturation, respiratory rate, and GCS to day 30 (Supplementary Fig. 4).

## Discussion

We present a comprehensive evaluation of a large retrospective cohort of COVID-19 patients from five Central Texas hospitals across SARS-CoV-2 waves between March 2020 and September 2022. We identified six distinct clinical trajectories that distinguished COVID-19 patient outcomes and provide a detailed analysis of risk factors and clinical features of COVID-19 disease severity in acute care patients. Interestingly, despite known prevalence of cardiopulmonary complications in COVID19 patients, our analysis reveals a multi-system failure in severe COVID-19 with a high association with neuropsychiatric co-morbidities and malnutrition.

Our cohort was similar to other studies in terms of median age and sex distribution.^3,18,19^ The overall 30-day mortality rate was 3.23%, with admitted patients at 7.73% and severe/critical TGs at 17.96%, in line with reported 11-23% rates.^20–22^ Similarly to other studies,^18,19^ we identified older age as a mortality risk factor, while younger age was associated with faster hospital discharge. Interestingly, the median age was similar between non-fatal severe/critical groups, suggesting that age may be a coarse risk factor for disease severity. Furthermore, males were over-represented in the most critical TGs (TG3/TG4), in line with sex as a risk factor of COVID-19 severity.^23,24^ Additionally, our results support Hispanic ethnicity correlating with severity.^25,26^ Notably, Hispanics formed a higher percentage of TG3, with higher rates of obesity and cardiovascular/renal comorbidities. This suggests that differences in outcomes among Hispanic patients may be associated with pre-existing chronic conditions, although some studies have attributed these disparities to delayed clinical presentation with more advanced disease.^27^

Over 2.5 years, there was a reduction in severe/critical cases within later COVID-19 waves. This reduction may be due to increased vaccinations by the end of Wave 2, less severe strains (Omicron), increasing immunity from previous infections, or increasing incidental hospital COVID-19 diagnoses. Vaccinations were most prevalent in TG1A patients, demonstrating the protective benefit against severe disease and mortality.^28^ However, further research is needed to ascertain whether vaccinations or less severe strains may account for lower severity with later waves.^29,30^

We observed a high frequency of neuropsychiatric diagnoses, including anxiety, depression, bipolar disorder, encephalopathy, dementia, and seizures and higher rates of central nervous system imaging in association with severe COVID-19 trajectories. These findings are in line with other studies finding higher neuropsychiatric complications in severe COVID-19.^31^ Despite the increase in case numbers in wave 4 (corresponding to the Omicron SARS-CoV-2 variant), the relative proportion of severe/critical COVID-19 patients receiving neurological and psychiatric consultations remained constant, indicating that neuropsychiatric complications persisted across different SARS-CoV-2 strains and did not abate with milder variants. Interestingly, the presence of neuropsychiatric diagnoses did not differentiate patients between critical trajectory groups (TG3 vs TG2 and TG4 vs TG3), suggesting that these diagnoses may have a lesser impact on COVID-19 related mortality compared to systemic diagnoses such as cardiac and respiratory complications. Nevertheless, TG3 patients were more likely to have brain imaging than TG2 patients, suggesting that concern of neurological complications remained high for patients with more severe COVID-19. Importantly, the observed higher association with neuropsychiatric complications in severe COVID-19 may inform the persistence of neuropsychiatric sequelae in Long-COVID patients following acute SARS-CoV-2 infection.^32^

One of the best severity-predictive lab markers at admission and throughout hospitalization was albumin, with the lowest level in TG4 and highest in TG1A. Although hypoalbuminemia is known to be a marker of infection, inflammation, acute and chronic illness,^33^ likely due to reduced liver synthesis, increased catabolism, or vascular permeability,^34^ albumin is not part of current COVID-19 risk-stratification algorithms. Based on our data, the inclusion of albumin in risk assessment could greatly improve prediction accuracy of COVID-19 outcomes. Additionally, while not the best marker of nutritional status,^35^ in association with higher malnutrition codes in severe/fatal patients, low albumin points to nutritional insufficiency as another key component in COVID-19 outcomes. Notably, other nutritional components, e.g. thiamine, Vitamin D, and B vitamins, which are also vital for neurological health, were not routinely checked in most patients, and multivitamins were administered to at most 20% of any TG, and only 10% of fatal patients. Our findings highlight the complex interaction between nutritional status and inflammation that is reflected by low albumin levels in severe patients and raises the question of whether evaluation of micronutrients, such as albumin, pre-albumin, thiamine, Vitamin D, and B vitamins, at initial clinical presentation in acute care patients, may help identify vulnerable patients, as well as whether measurement and replacement of micronutrients may benefit patient outcomes.

Acid-base dysregulation was another set of diagnoses enriched in severe TGs. Notably, anion gap, a measure of acid-base homeostasis, increased in severe TGs in association with low albumin. The high anion gap may be explained by decline in renal function or higher levels of sepsis contributing to lactic acidosis in severe patients. Physiologically, acid base dysregulation can lead to abnormalities across multiple organs but notably within the cardiovascular system. Indeed, cardiac dysfunction increased across severe/critical TGs, with highest rates in the fatal TG4.^36^ Given more severe outcomes in patients with acid base disorders, it is critical for physicians to promptly identify and correct acid-base dysregulation causes toward improving COVID-19 outcomes.

Immunologically, our results support association of high neutrophils and immature granulocytes and low lymphocytes and monocytes in severe patients.^37^ Notably, we did not observe a significant difference between TG3 and TG4 at admission, suggesting that these immune markers may be reflective of severity and not mortality. We also observed higher rates of sepsis, shock, and immunodeficiency in critical patients, with RA and IBD over-represented in critical TGs, supporting evidence that certain immune conditions may predispose to more severe COVID-19.^38^

There were several limitations to our study. First, our cohort was from a single health system, and was disproportionately Hispanic and white. Second, all data were retrieved from EHRs, known to have missing data and lower quality data with respect to demographics, e.g. race and ethnicity. ^39^ Also, our clinical dataset did not include viral load or SARS-CoV-2 variant information, though we were able to use epidemiological data to estimate the prevalence of strains over time.

In conclusion, our identified clinical signatures could help clinicians better predict disease trajectories in acute COVID-19 and anticipate elevation of care versus discharge. Importantly, we highlight an association between COVID-19 severity and neuropsychiatric complications as well as nutritional insufficiency as key risk factors for COVID-19 outcomes, raising the urgency for prompt evaluation and treatment of these conditions toward improving patient outcomes in the ED, hospital and post discharge.

## Acknowledgements

We thank the COVID-19 patients of Central Texas Ascension Seton medical system for participating in this study. We thank Dr. William Schwartz for thoughtful comments on the manuscript and appreciate discussions with Dr. Greg Wallingford, Dr. Lauren Ehrlich and Nadia Siles. We appreciate Dr. Lauren Ehrlich sharing computational servers from Advance Microdevices and participating in obtaining study funding. We would like to thank Kristen Crabtree for advice and assistance with institutional review board evaluation of our study, as well as Patrick Boswell for help with data transfer. We are also grateful for the administrative and technical support from Dell Medical School Neurology Department and the UT Austin Biomedical Resource Computational Facility.

