## supplemental_material for "Dissecting Clinical Features of COVID-19 in a Cohort of 21,312 Acute Care Patients"

#### Supplemental Methods

##### Unsupervised Latent Class Mixed Modeling Extended

Unsupervised latent class mixed modeling was used to identify patient clinical trajectories for the modified WHO ordinal score over the course of 30 days. Quadratic models were calculated ranging from 1-7 latent groups with either no random, a linear time random, or a quadratic time random variable, and similarly Cubic models were calculated for 1-7 latent groups with a linear time random or no random variable. Best fit was achieved by the four-group quadratic time random model based on a combination of Bayesian Information Criteria (BIC) and entropy (Supplemental Figure 1A and 1B). TG3 contained 23 patients who died after day 26 which we moved to TG4 to create a 'death by day 30' grouping and moved the 10 patients from TG4 who did not die by day 30 into TG3. TG1 was the largest patient group and had the fastest hospital discharge. Due to the sizable population of this group (n=17,476) and to further distinguish acute hospitalized trajectories, we repeated latent class mixed modeling with TG1 patients over a 12-day period, resulting in the identification of three subclasses: TG1A, TG1B, and TG1C (Supplemental Figure 1C, 1D, and 2).

##### Supplemental Table Generation of Frequency of Diagnoses, Consultations, Imaging, Administered Medications, Laboratory Tests, and Vital Signs

The frequency of diagnoses, consultations, and administered medications that at least 10% of any TG received are reported in these tables. For diagnoses, consultations, and administered medications, some additional features of particular interest were added that occurred in less than 10% of the TG including neuropsychiatric and immune diagnoses of interest. For administered medications, patients were included based on receiving the medication at least once during length of stay. Dosing or frequency of administration were not evaluated. For imaging, any imaging that occurred in greater than 5 % of a TG is reported. The ‘Frequency’ column graphs the relative frequency between the TGs reported as a % in the prior columns (i.e. for each TG what percent of patients was positive). The ‘Distribution’ column graphs the proportion of TG among patients with that respective feature (i.e. for positive patients what was the TG composition). Bonferroni adjusted p-values are reported in the final heatstrip column, with color indicating p.adj <= 0.05. For the global p.value, chi-square testing was used for all groups, where pairwise columns focused on the specific comparison using chi-square testing with all p-values reported Bonferroni adjusted. For the Diagnoses and Consultations identified in Table S4 and S5, they were also evaluated in total frequency between the five COVID-19 waves identified which are reported in Table S2 and S3.

##### Cumulative Linked Modeling

For modeling of laboratory test and vital signs on or near hospital admission (day 0-1), the R package ordinal (v2022.11.16) was used to evaluate differences between TGs while controlling for hospital site, year/yearly quarter, sex, age, ethnicity, and race. Model results are reported in Table S8 and S9.

##### Generalized Additive Mixed Modeling

For longitudinal modeling of laboratory tests and vital signs, the R package (gamm4) (v0.2.6) was used to evaluate differences between TGs while controlling for hospital site, year/yearly quarter, sex, age, ethnicity, and race with patient as a mixed effect. If patients had multiple measures on the same day their average was used. Model results are reported in Table S10 and S11 and graphed in Figure S3 and S4, with TG drop out when less than 10% of each group had measurements.

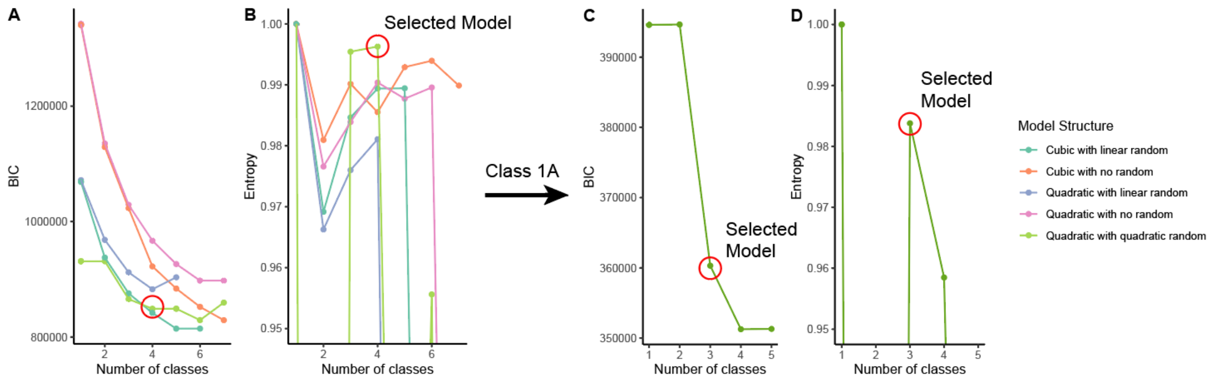


**Figure S1 - Latent Class Mixed Modeling yielded the highest Entropy model from a Quadratic model with quadratic time random variable. A)** Bayesian Information Criteria (BIC) and **B)** Entropy of the evaluated models structures for modeling the modified WHO score over 30 days. **C)** Bayesian Information Criteria (BIC) and **D)** Entropy of the evaluated models structures for modeling the modified WHO score over 30 days to identify subclasses of TG1.

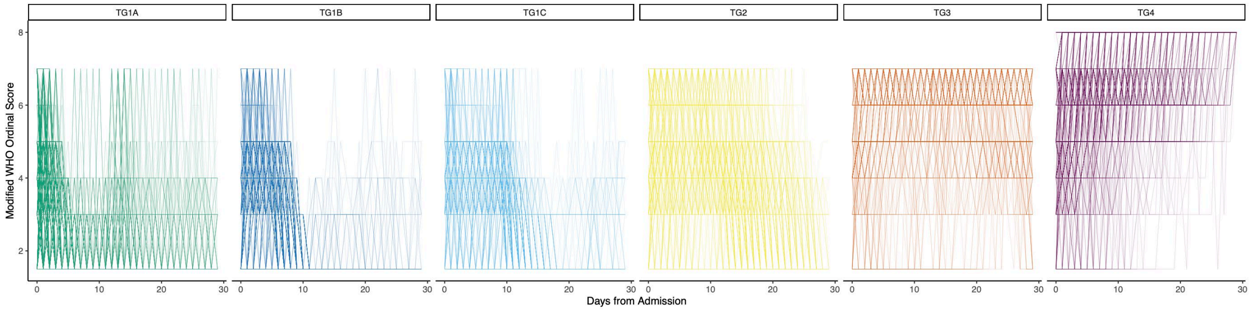


**Figure S2 -** Noodle plot of individual patients in each trajectory group.

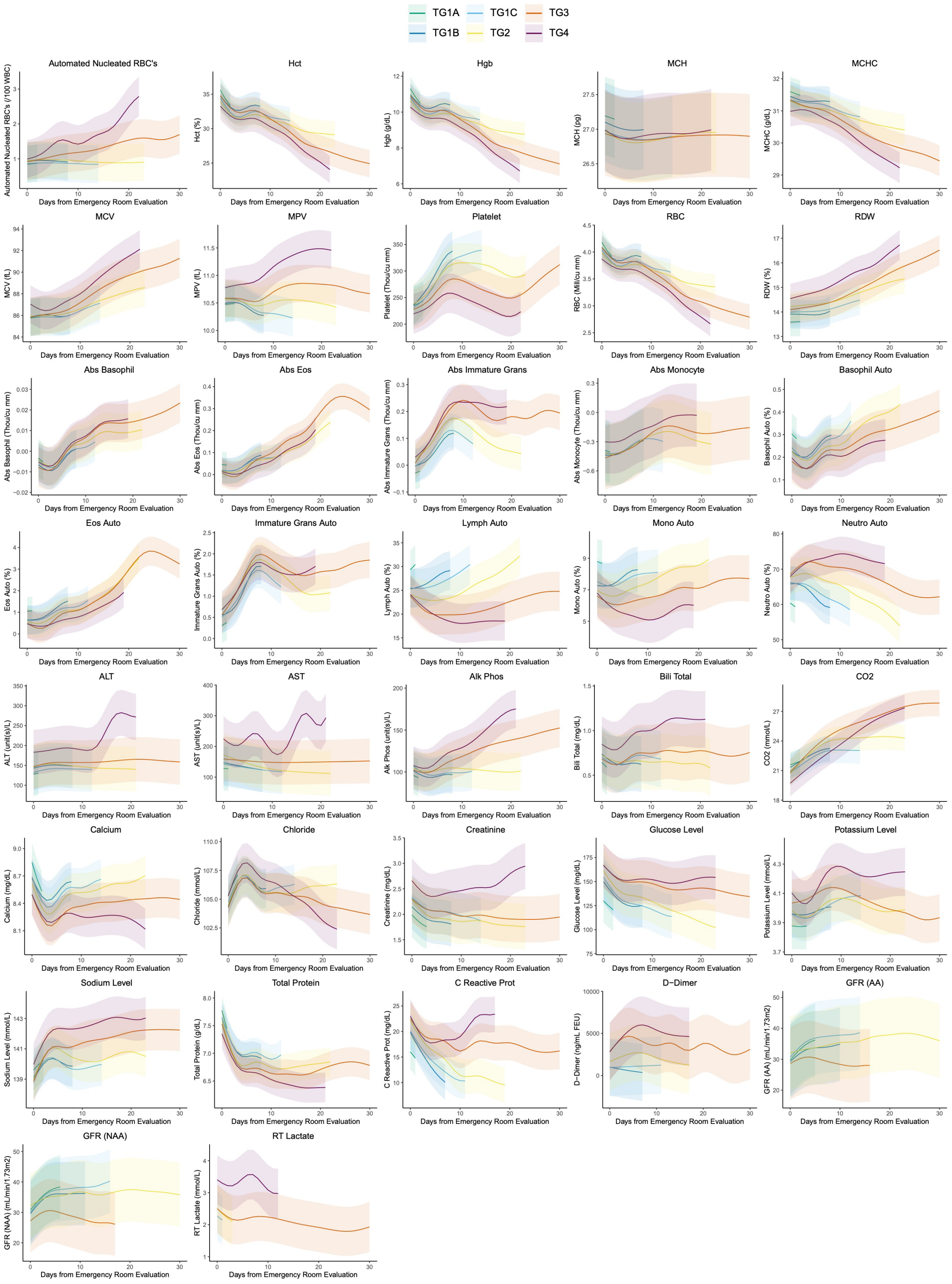


**Figure S3 –** Additional Laboratory Tests across Acute Care by Trajectory Group. Shaded region denotes 95% confidence intervals for each Trajectory Group. Laboratory testing modeling results by TG were trimmed to days with data for greater than 10% of the TG.

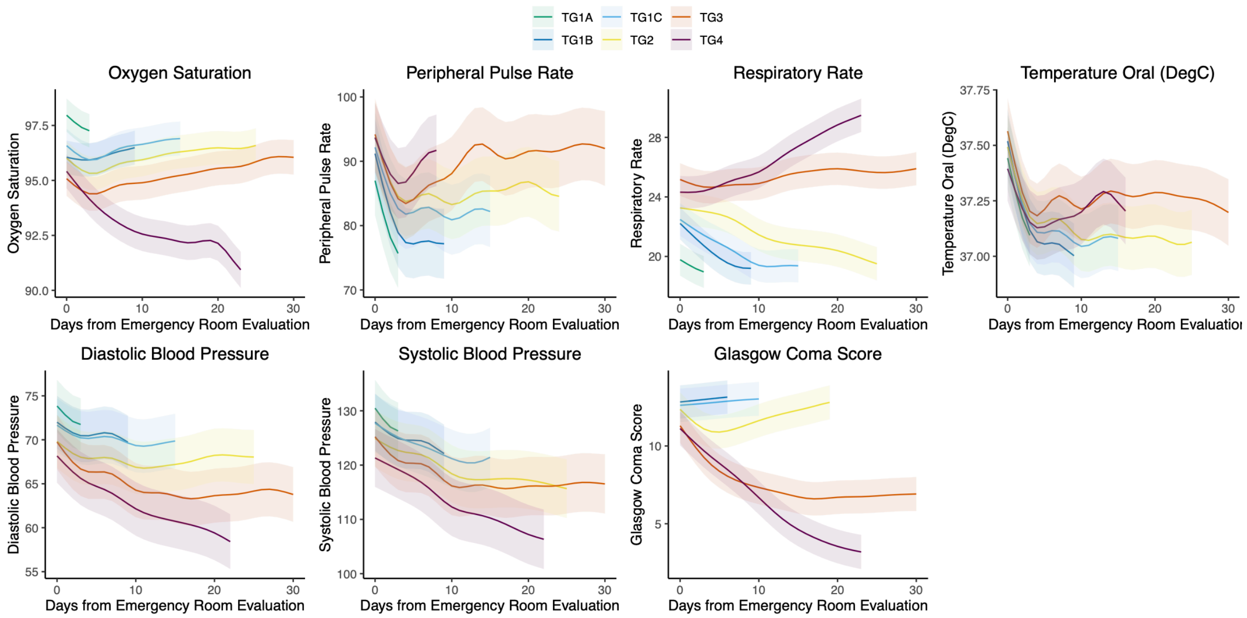


**Figure S4 -** Vital Signs across Acute Care by Trajectory Group. Shaded region denotes 95% confidence intervals for each Trajectory Group. Laboratory testing modeling results by TG were trimmed to days with data for greater than 10% of the TG.

### Table S2 - Diagnoses by COVID-19 Wave

| Diagnoses | Total | Alpha/Beta: Wave 1 (03-01-2020 to 10-01-2020) | Alpha/Beta: Wave 2 (10-01-2020 to 06-15-2021) | Delta: Wave 3 (06-15-2021 to 11-15-2021) | Omicron: Wave 4 (11-15-2021 to 04-01-2022) | Omicron: Wave 5 (04-01-2022 to 09-01-2022) |
|---|---|---|---|---|---|---|
| Acidosis | 5.607% (1195) | 8.98% (295) | 6.754% (362) | 6.135% (263) | 3.335% (196) | 3.156% (79) |
| Actively Intoxicated | 0.511% (109) | 0.457% (15) | 0.597% (32) | 0.373% (16) | 0.459% (27) | 0.759% (19) |
| Acute and chronic respiratory failure with hypoxia | 1.206% (257) | 1.431% (47) | 1.362% (73) | 1.213% (52) | 0.953% (56) | 1.159% (29) |
| Acute kidney failure | 10.098% (2152) | 14.886% (489) | 12.146% (651) | 10.404% (446) | 6.976% (410) | 6.233% (156) |
| Acute posthemorrhagic anemia | 1.412% (301) | 1.887% (62) | 1.791% (96) | 1.19% (51) | 1.259% (74) | 0.719% (18) |
| Acute respiratory distress | 19.576% (4172) | 30.472% (1001) | 26.959% (1445) | 25.122% (1077) | 8.678% (510) | 5.553% (139) |
| Adjustment disorder with depressed mood | 0.042% (9) | 0.061% (2) | 0.037% (2) | 0.093% (4) | 0% (0) | 0.04% (1) |
| Adjustment disorders | 0.131% (28) | 0.152% (5) | 0.112% (6) | 0.28% (12) | 0.034% (2) | 0.12% (3) |
| Adverse effect of glucocorticoids and synthetic analogues, initial encounter | 3.735% (796) | 4.566% (150) | 5.541% (297) | 4.852% (208) | 1.77% (104) | 1.478% (37) |
| Alcohol Abuse | 1.905% (406) | 1.979% (65) | 1.903% (102) | 1.633% (70) | 1.957% (115) | 2.157% (54) |
| Alcohol related disorders | 1.905% (406) | 1.979% (65) | 1.903% (102) | 1.633% (70) | 1.957% (115) | 2.157% (54) |
| Alkalosis | 0.981% (209) | 1.918% (63) | 1.362% (73) | 1.003% (43) | 0.425% (25) | 0.2% (5) |
| Alzheimer's disease | 0.338% (72) | 0.639% (21) | 0.448% (24) | 0.14% (6) | 0.255% (15) | 0.24% (6) |
| Amnestic disorder due to known physiological condition | 0.005% (1) | 0% (0) | 0% (0) | 0% (0) | 0.017% (1) | 0% (0) |
| Anemia | 5.856% (1248) | 7.61% (250) | 6.381% (342) | 6.018% (258) | 4.764% (280) | 4.714% (118) |
| Ankylosing spondylitis | 0.028% (6) | 0% (0) | 0.037% (2) | 0.023% (1) | 0.051% (3) | 0% (0) |
| Anorexia nervosa | 0.009% (2) | 0% (0) | 0% (0) | 0.023% (1) | 0.017% (1) | 0% (0) |
| Antisocial personality disorder | 0.014% (3) | 0% (0) | 0.019% (1) | 0.047% (2) | 0% (0) | 0% (0) |
| Anxiety disorder, unspecified | 6.691% (1426) | 7.458% (245) | 8.06% (432) | 7.884% (338) | 4.917% (289) | 4.874% (122) |
| Asthma | 5.335% (1137) | 6.119% (201) | 5.877% (315) | 5.668% (243) | 5.377% (316) | 2.477% (62) |
| Athereosclretic heart disease | 5.861% (1249) | 7.397% (243) | 7.481% (401) | 4.619% (198) | 5.173% (304) | 4.115% (103) |
| Attention-deficit hyperactivity disorders | 0.343% (73) | 0.152% (5) | 0.299% (16) | 0.56% (24) | 0.357% (21) | 0.28% (7) |
| Autistic disorder | 0.075% (16) | 0.091% (3) | 0.056% (3) | 0.093% (4) | 0.085% (5) | 0.04% (1) |
| Autoimmune Hepatitis | 0.033% (7) | 0.061% (2) | 0% (0) | 0.047% (2) | 0.051% (3) | 0% (0) |
| Bell's palsy | 0.089% (19) | 0.122% (4) | 0.131% (7) | 0.047% (2) | 0.085% (5) | 0.04% (1) |
| Benign intracranial hypertension | 0.033% (7) | 0.03% (1) | 0.019% (1) | 0.023% (1) | 0.068% (4) | 0% (0) |
| Binge eating disorder | 0.005% (1) | 0.03% (1) | 0% (0) | 0% (0) | 0% (0) | 0% (0) |
| Bipolar | 1.68% (358) | 1.887% (62) | 1.978% (106) | 1.656% (71) | 1.344% (79) | 1.598% (40) |
| Bipolar disorder | 1.68% (358) | 1.887% (62) | 1.978% (106) | 1.656% (71) | 1.344% (79) | 1.598% (40) |
| Borderline intellectual functioning | 0.005% (1) | 0.03% (1) | 0% (0) | 0% (0) | 0% (0) | 0% (0) |
| Borderline personality disorder | 0.122% (26) | 0.091% (3) | 0.149% (8) | 0.093% (4) | 0.119% (7) | 0.16% (4) |
| Brief psychotic disorder | 0.122% (26) | 0.03% (1) | 0.187% (10) | 0.07% (3) | 0.102% (6) | 0.24% (6) |
| Burn-out | 4.833% (1030) | 0% (0) | 11.026% (591) | 8.071% (346) | 1.191% (70) | 0.919% (23) |
| Cannabis related disorders | 0.859% (183) | 0.944% (31) | 0.728% (39) | 0.886% (38) | 0.97% (57) | 0.719% (18) |
| Cardiac arrest, cause unspecified | 0.995% (212) | 1.187% (39) | 1.25% (67) | 1.54% (66) | 0.579% (34) | 0.24% (6) |
| Celiac disease | 0.033% (7) | 0.061% (2) | 0.056% (3) | 0% (0) | 0.017% (1) | 0.04% (1) |
| Celiac Disease | 0.033% (7) | 0.061% (2) | 0.056% (3) | 0% (0) | 0.017% (1) | 0.04% (1) |
| Cerebral infarction | 0.957% (204) | 1.035% (34) | 1.138% (61) | 0.863% (37) | 0.97% (57) | 0.599% (15) |
| Cerebral palsy | 0.141% (30) | 0.244% (8) | 0.187% (10) | 0.07% (3) | 0.085% (5) | 0.16% (4) |
| Cerebral palsy and other paralytic syndromes | 0.948% (202) | 1.218% (40) | 0.933% (50) | 1.003% (43) | 0.783% (46) | 0.919% (23) |
| Chronic inflammatory demyelinating polyneuritis | 0.033% (7) | 0.03% (1) | 0.075% (4) | 0.023% (1) | 0.017% (1) | 0% (0) |
| Chronic Kidney Disease | 3.918% (835) | 6.697% (220) | 5.019% (269) | 2.776% (119) | 2.603% (153) | 2.956% (74) |
| Chronic obstructive pulmonary disease with (acute) lower respiratory infection | 1.567% (334) | 1.857% (61) | 2.127% (114) | 1.353% (58) | 1.191% (70) | 1.239% (31) |
| Chronic pain syndrome | 0.216% (46) | 0.274% (9) | 0.205% (11) | 0.187% (8) | 0.17% (10) | 0.32% (8) |
| Cluster headaches and other trigeminal autonomic cephalgias (TAC) | 0.009% (2) | 0% (0) | 0.019% (1) | 0% (0) | 0% (0) | 0.04% (1) |
| Cocaine related disorders | 0.469% (100) | 0.609% (20) | 0.522% (28) | 0.42% (18) | 0.425% (25) | 0.36% (9) |
| Conduct disorder, unspecified | 0.019% (4) | 0.03% (1) | 0.037% (2) | 0% (0) | 0.017% (1) | 0% (0) |
| Conduct disorders | 0.042% (9) | 0.03% (1) | 0.056% (3) | 0.047% (2) | 0.051% (3) | 0% (0) |
| Constipation | 3.308% (705) | 4.992% (164) | 3.769% (202) | 3.662% (157) | 2.28% (134) | 1.918% (48) |
| Conversion disorder with motor symptom or deficit | 0.005% (1) | 0% (0) | 0% (0) | 0.023% (1) | 0% (0) | 0% (0) |
| COPD | 3.12% (665) | 3.196% (105) | 3.545% (190) | 2.309% (99) | 3.216% (189) | 3.276% (82) |
| Cough | 16.094% (3430) | 16.773% (551) | 15.746% (844) | 21.04% (902) | 12.217% (718) | 16.58% (415) |
| Degenerative disease of nervous system, unspecified | 0.033% (7) | 0.03% (1) | 0.075% (4) | 0% (0) | 0.034% (2) | 0% (0) |
| Dehydration | 5.748% (1225) | 7.275% (239) | 6.53% (350) | 5.785% (248) | 4.22% (248) | 5.593% (140) |
| Delirium due to known physiological condition | 0.521% (111) | 0.974% (32) | 0.69% (37) | 0.373% (16) | 0.357% (21) | 0.2% (5) |
| Delusional disorders | 0.183% (39) | 0.122% (4) | 0.131% (7) | 0.233% (10) | 0.221% (13) | 0.2% (5) |
| Dependence on respirator [ventilator] status | 0.338% (72) | 0.396% (13) | 0.336% (18) | 0.606% (26) | 0.221% (13) | 0.08% (2) |
| Dependent personality disorder | 0.005% (1) | 0% (0) | 0% (0) | 0.023% (1) | 0% (0) | 0% (0) |
| Depression | 4.978% (1061) | 5.662% (186) | 6.231% (334) | 4.129% (177) | 4.713% (277) | 3.476% (87) |
| Depressive episode | 4.978% (1061) | 5.662% (186) | 6.231% (334) | 4.129% (177) | 4.713% (277) | 3.476% (87) |
| Disappearance and death of family member | 0.08% (17) | 0.122% (4) | 0.019% (1) | 0.093% (4) | 0.034% (2) | 0.24% (6) |
| Disorders of calcium metabolism | 0.967% (206) | 1.218% (40) | 1.231% (66) | 0.956% (41) | 0.749% (44) | 0.599% (15) |
| Disorders of copper metabolism | 0.005% (1) | 0% (0) | 0% (0) | 0% (0) | 0% (0) | 0.04% (1) |
| Disorders of phosphorus metabolism and phosphatases | 1.154% (246) | 2.253% (74) | 1.343% (72) | 0.77% (33) | 0.868% (51) | 0.639% (16) |
| Dissociative and conversion disorder, unspecified | 0.033% (7) | 0% (0) | 0.075% (4) | 0.023% (1) | 0.034% (2) | 0% (0) |
| Dissociative identity disorder | 0.009% (2) | 0% (0) | 0% (0) | 0.023% (1) | 0.017% (1) | 0% (0) |
| Do not resuscitate | 6.203% (1322) | 8.371% (275) | 7.854% (421) | 6.928% (297) | 3.965% (233) | 3.835% (96) |
| Dysphagia, unspecified | 1.553% (331) | 2.222% (73) | 1.623% (87) | 1.003% (43) | 1.48% (87) | 1.638% (41) |
| Dysthymic disorder | 0.023% (5) | 0% (0) | 0.037% (2) | 0% (0) | 0.034% (2) | 0.04% (1) |
| Elevated blood-pressure | 0.572% (122) | 0.944% (31) | 0.672% (36) | 0.42% (18) | 0.374% (22) | 0.599% (15) |
| Encephalitis, myelitis and encephalomyelitis, unspecified | 0.047% (10) | 0.061% (2) | 0.037% (2) | 0.047% (2) | 0.068% (4) | 0% (0) |
| Encephalopathy | 3.205% (683) | 4.17% (137) | 3.806% (204) | 2.916% (125) | 2.739% (161) | 2.237% (56) |
| Encounter for palliative care | 4.57% (974) | 6.575% (216) | 5.093% (273) | 6.345% (272) | 2.859% (168) | 1.798% (45) |
| Epilepsy and recurrent seizures | 1.45% (309) | 1.918% (63) | 1.828% (98) | 1.143% (49) | 1.31% (77) | 0.879% (22) |
| Facial nerve disorders | 0.089% (19) | 0.122% (4) | 0.131% (7) | 0.047% (2) | 0.085% (5) | 0.04% (1) |
| Fatty (change of) liver, not elsewhere classified | 0.746% (159) | 1.218% (40) | 0.821% (44) | 0.676% (29) | 0.63% (37) | 0.36% (9) |
| Fever, unspecified | 9.896% (2109) | 12.633% (415) | 9.123% (489) | 12.363% (530) | 6.636% (390) | 11.386% (285) |
| Fluid overload, unspecified | 1.178% (251) | 1.644% (54) | 1.642% (88) | 0.956% (41) | 0.902% (53) | 0.599% (15) |
| Gastro-esophageal reflux disease without esophagitis | 4.481% (955) | 5.54% (182) | 5.877% (315) | 3.802% (163) | 3.607% (212) | 3.316% (83) |
| Gender identity disorder, unspecified | 0.028% (6) | 0.03% (1) | 0.075% (4) | 0.023% (1) | 0% (0) | 0% (0) |
| Gender identity disorders | 0.028% (6) | 0.03% (1) | 0.075% (4) | 0.023% (1) | 0% (0) | 0% (0) |
| Generalized anxiety disorder | 0.385% (82) | 0.761% (25) | 0.485% (26) | 0.187% (8) | 0.272% (16) | 0.28% (7) |
| Gout | 0.882% (188) | 0.883% (29) | 1.101% (59) | 1.05% (45) | 0.664% (39) | 0.639% (16) |
| Grave's Disease | 0.047% (10) | 0.061% (2) | 0.037% (2) | 0.023% (1) | 0.051% (3) | 0.08% (2) |
| Guillain-Barre syndrome | 0.033% (7) | 0.03% (1) | 0.056% (3) | 0.023% (1) | 0.034% (2) | 0% (0) |
| Hallucinogen related disorders | 0.047% (10) | 0.03% (1) | 0.019% (1) | 0.047% (2) | 0.085% (5) | 0.04% (1) |
| Hashimoto's Throiditis | 0.108% (23) | 0.061% (2) | 0.112% (6) | 0.21% (9) | 0.051% (3) | 0.12% (3) |
| Headache | 4.096% (873) | 3.714% (122) | 3.825% (205) | 3.872% (166) | 3.845% (226) | 6.153% (154) |
| Heart Failure | 5.748% (1225) | 6.758% (222) | 6.586% (353) | 4.549% (195) | 5.632% (331) | 4.954% (124) |
| Hereditary and idiopathic neuropathy, unspecified | 0.005% (1) | 0% (0) | 0% (0) | 0.023% (1) | 0% (0) | 0% (0) |
| Hereditary motor and sensory neuropathy | 0.009% (2) | 0.03% (1) | 0% (0) | 0% (0) | 0.017% (1) | 0% (0) |
| Huntington's disease | 0.009% (2) | 0.03% (1) | 0% (0) | 0% (0) | 0% (0) | 0.04% (1) |
| Hyperglycemia | 4.049% (863) | 5.175% (170) | 5.205% (279) | 5.085% (218) | 2.552% (150) | 1.838% (46) |
| Hyperglycemia, unspecified | 4.049% (863) | 5.175% (170) | 5.205% (279) | 5.085% (218) | 2.552% (150) | 1.838% (46) |
| Hyperkalemia | 3.463% (738) | 4.11% (135) | 4.067% (218) | 3.616% (155) | 2.757% (162) | 2.717% (68) |
| Hyperlipidemia, unspecified | 12.383% (2639) | 16.104% (529) | 16.903% (906) | 10.847% (465) | 9.41% (553) | 7.431% (186) |
| Hyperosmolality and hypernatremia | 2.501% (533) | 4.718% (155) | 3.041% (163) | 2.659% (114) | 1.344% (79) | 0.879% (22) |
| Hypertension | 21.978% (4684) | 28.919% (950) | 27.071% (1451) | 22.627% (970) | 17.458% (1026) | 11.466% (287) |
| Hypertensive heart disease with heart failure | 2.801% (597) | 2.953% (97) | 3.134% (168) | 2.309% (99) | 2.995% (176) | 2.277% (57) |
| Hypo-osmolality and hyponatremia | 7.306% (1557) | 11.872% (390) | 8.731% (468) | 7.768% (333) | 4.526% (266) | 3.995% (100) |
| Hypoglycemia, unspecified | 0.755% (161) | 0.974% (32) | 0.784% (42) | 0.886% (38) | 0.647% (38) | 0.439% (11) |
| Hypokalemia | 5.809% (1238) | 7.732% (254) | 7.649% (410) | 6.228% (267) | 3.931% (231) | 3.036% (76) |
| Hypomagnesemia | 1.014% (216) | 1.492% (49) | 0.896% (48) | 0.84% (36) | 0.919% (54) | 1.159% (29) |
| Hypotension | 1.811% (386) | 2.1% (69) | 2.276% (122) | 1.959% (84) | 1.106% (65) | 1.838% (46) |
| Hypothyroidism | 4.819% (1027) | 6.027% (198) | 6.138% (329) | 4.269% (183) | 4.033% (237) | 3.196% (80) |
| Hypoxemia | 9.516% (2028) | 15.282% (502) | 12.873% (690) | 12.223% (524) | 4.203% (247) | 2.597% (65) |
| Ileus, unspecified | 0.432% (92) | 0.487% (16) | 0.672% (36) | 0.373% (16) | 0.357% (21) | 0.12% (3) |
| Impulse disorder, unspecified | 0.009% (2) | 0.03% (1) | 0.019% (1) | 0% (0) | 0% (0) | 0% (0) |
| Imunnodeficiency | 0.629% (134) | 0.03% (1) | 0.728% (39) | 0.653% (28) | 0.868% (51) | 0.599% (15) |
| Inflammatory Bowel Disease | 0.948% (202) | 1.218% (40) | 1.101% (59) | 0.63% (27) | 0.936% (55) | 0.839% (21) |
| Insomnia not due to a substance or known physiological condition | 0.047% (10) | 0.091% (3) | 0.093% (5) | 0% (0) | 0.017% (1) | 0.04% (1) |
| Insulin Use | 6.658% (1419) | 10.015% (329) | 8.358% (448) | 6.368% (273) | 5.02% (295) | 2.956% (74) |
| Intellectual disabilities | 0.286% (61) | 0.609% (20) | 0.448% (24) | 0.187% (8) | 0.085% (5) | 0.16% (4) |
| Intermittent explosive disorder | 0.009% (2) | 0.03% (1) | 0% (0) | 0% (0) | 0% (0) | 0.04% (1) |
| Intracranial abscess and granuloma | 0.005% (1) | 0.03% (1) | 0% (0) | 0% (0) | 0% (0) | 0% (0) |
| Iron deficiency anemia, unspecified | 1.633% (348) | 2.405% (79) | 1.903% (102) | 1.12% (48) | 1.548% (91) | 1.119% (28) |
| Liver disease | 1.136% (242) | 4.292% (141) | 0.914% (49) | 0.606% (26) | 0.374% (22) | 0.16% (4) |
| Long term (current) use of oral hypoglycemic drugs | 4.974% (1060) | 7.976% (262) | 5.933% (318) | 3.895% (167) | 4.356% (256) | 2.277% (57) |
| Long term asprin | 6.405% (1365) | 7.58% (249) | 7.873% (422) | 4.945% (212) | 6.228% (366) | 4.634% (116) |
| Long Term Coagulants | 3.96% (844) | 3.531% (116) | 4.44% (238) | 4.315% (185) | 3.539% (208) | 3.875% (97) |
| Long term drug therapy | 17.624% (3756) | 19.33% (635) | 19.216% (1030) | 16.025% (687) | 17.594% (1034) | 14.782% (370) |
| Lupus | 0.347% (74) | 0.244% (8) | 0.41% (22) | 0.397% (17) | 0.357% (21) | 0.24% (6) |
| Magnesium deficiency | 0.009% (2) | 0% (0) | 0% (0) | 0.023% (1) | 0.017% (1) | 0% (0) |
| Malignant neoplasm of brain, unspecified | 0.014% (3) | 0% (0) | 0% (0) | 0% (0) | 0.034% (2) | 0.04% (1) |
| Malingerer [conscious simulation] | 0.033% (7) | 0.061% (2) | 0% (0) | 0% (0) | 0.051% (3) | 0.08% (2) |
| Malnutrition | 2.102% (448) | 2.557% (84) | 2.239% (120) | 1.889% (81) | 1.957% (115) | 1.918% (48) |
| Migraine | 0.99% (211) | 0.883% (29) | 1.399% (75) | 0.863% (37) | 0.919% (54) | 0.639% (16) |
| Mixed disorder of acid-base balance | 1.248% (266) | 2.77% (91) | 1.418% (76) | 1.423% (61) | 0.562% (33) | 0.2% (5) |
| Moderate protein-calorie malnutrition | 0.371% (79) | 0.365% (12) | 0.354% (19) | 0.327% (14) | 0.374% (22) | 0.479% (12) |
| Mood disorder due to known physiological condition | 0.014% (3) | 0% (0) | 0.019% (1) | 0.023% (1) | 0% (0) | 0.04% (1) |
| Motor neuron disease | 0.009% (2) | 0% (0) | 0% (0) | 0% (0) | 0.017% (1) | 0.04% (1) |
| Multiple Sclerosis | 0.136% (29) | 0.122% (4) | 0.187% (10) | 0.117% (5) | 0.119% (7) | 0.12% (3) |
| Myasthenia gravis | 0.038% (8) | 0.061% (2) | 0.056% (3) | 0.023% (1) | 0.034% (2) | 0% (0) |
| Myoclonus | 0.103% (22) | 0.091% (3) | 0.075% (4) | 0.14% (6) | 0.153% (9) | 0% (0) |
| Narcolepsy and cataplexy | 0.014% (3) | 0.03% (1) | 0.019% (1) | 0.023% (1) | 0% (0) | 0% (0) |
| Neurocognitive disorder with Lewy bodies | 0.056% (12) | 0.091% (3) | 0.056% (3) | 0.023% (1) | 0.068% (4) | 0.04% (1) |
| Neuromyelitis optica [Devic] | 0.009% (2) | 0% (0) | 0% (0) | 0% (0) | 0.034% (2) | 0% (0) |
| Niacin deficiency [pellagra] | 0.005% (1) | 0% (0) | 0.019% (1) | 0% (0) | 0% (0) | 0% (0) |
| Nictoine Dependence | 6.808% (1451) | 9.315% (306) | 8.563% (459) | 5.575% (239) | 5.275% (310) | 5.473% (137) |
| Nontraumatic intracerebral hemorrhage | 0.249% (53) | 0.213% (7) | 0.354% (19) | 0.28% (12) | 0.238% (14) | 0.04% (1) |
| Obesity | 8.418% (1794) | 11.385% (374) | 10.69% (573) | 10.17% (436) | 5.411% (318) | 3.716% (93) |
| Obesity due to excess calories | 3.5% (746) | 4.81% (158) | 4.552% (244) | 4.525% (194) | 1.974% (116) | 1.358% (34) |
| Obesity, unspecified | 5.213% (1111) | 6.88% (226) | 6.455% (346) | 6.228% (267) | 3.624% (213) | 2.357% (59) |
| Obsessive-compulsive disorder | 0.075% (16) | 0.061% (2) | 0.075% (4) | 0.093% (4) | 0.085% (5) | 0.04% (1) |
| Obsessive-compulsive disorder, unspecified | 0.066% (14) | 0.061% (2) | 0.075% (4) | 0.047% (2) | 0.085% (5) | 0.04% (1) |
| Obstructive sleep apnea | 2.599% (554) | 3.288% (108) | 3.731% (200) | 2.263% (97) | 1.855% (109) | 1.598% (40) |
| Opioid related disorders | 0.38% (81) | 0.304% (10) | 0.504% (27) | 0.35% (15) | 0.357% (21) | 0.32% (8) |
| Other abnormalities of plasma proteins | 0.957% (204) | 0% (0) | 0.97% (52) | 1.376% (59) | 1.191% (70) | 0.919% (23) |
| Other disorders of electrolyte and fluid balance, not elsewhere classified | 0.704% (150) | 0.791% (26) | 0.821% (44) | 0.91% (39) | 0.425% (25) | 0.639% (16) |
| Other disorders of phosphorus metabolism | 1.154% (246) | 2.253% (74) | 1.343% (72) | 0.77% (33) | 0.868% (51) | 0.639% (16) |
| Other eating disorders | 0.009% (2) | 0.03% (1) | 0.019% (1) | 0% (0) | 0% (0) | 0% (0) |
| Other encephalitis and encephalomyelitis | 0.028% (6) | 0.061% (2) | 0.019% (1) | 0.047% (2) | 0% (0) | 0.04% (1) |
| Other inflammatory polyneuropathies | 0.038% (8) | 0.03% (1) | 0.093% (5) | 0.023% (1) | 0.017% (1) | 0% (0) |
| Other specified abnormal findings of blood chemistry | 2.886% (615) | 4.749% (156) | 4.422% (237) | 2.309% (99) | 1.548% (91) | 1.278% (32) |
| Other specified abnormalities of plasma proteins | 0.957% (204) | 0% (0) | 0.97% (52) | 1.376% (59) | 1.191% (70) | 0.919% (23) |
| Other specified anxiety disorders | 0.108% (23) | 0.152% (5) | 0.112% (6) | 0.07% (3) | 0.119% (7) | 0.08% (2) |
| Other specified eating disorder | 0.005% (1) | 0% (0) | 0.019% (1) | 0% (0) | 0% (0) | 0% (0) |
| Other specified metabolic disorders | 0.122% (26) | 0.213% (7) | 0.168% (9) | 0.023% (1) | 0.119% (7) | 0.08% (2) |
| Other sphingolipidosis | 0.005% (1) | 0% (0) | 0% (0) | 0.023% (1) | 0% (0) | 0% (0) |
| Other vitamin deficiencies | 0.019% (4) | 0.061% (2) | 0% (0) | 0% (0) | 0.017% (1) | 0.04% (1) |
| Panic disorder [episodic paroxysmal anxiety] | 0.399% (85) | 0.183% (6) | 0.578% (31) | 0.606% (26) | 0.255% (15) | 0.28% (7) |
| Paranoid personality disorder | 0.014% (3) | 0% (0) | 0.019% (1) | 0% (0) | 0.034% (2) | 0% (0) |
| Parkinson's disease | 0.436% (93) | 0.487% (16) | 0.522% (28) | 0.303% (13) | 0.306% (18) | 0.719% (18) |
| Phobic anxiety disorders | 0.033% (7) | 0% (0) | 0.037% (2) | 0% (0) | 0.068% (4) | 0.04% (1) |
| Pneumonia | 29.087% (6199) | 46.514% (1528) | 38.526% (2065) | 38.558% (1653) | 12.932% (760) | 7.711% (193) |
| Polyarteritis Nodosa | 0.019% (4) | 0.03% (1) | 0.037% (2) | 0% (0) | 0.017% (1) | 0% (0) |
| Post-traumatic stress disorder (PTSD) | 0.784% (167) | 0.578% (19) | 0.914% (49) | 0.84% (36) | 0.783% (46) | 0.679% (17) |
| Postsurgical malabsorption, not elsewhere classified | 0.005% (1) | 0% (0) | 0.019% (1) | 0% (0) | 0% (0) | 0% (0) |
| Prediabetes | 0.652% (139) | 0.913% (30) | 0.914% (49) | 0.793% (34) | 0.272% (16) | 0.4% (10) |
| Primary Biliary Cirrhosis | 0.019% (4) | 0% (0) | 0% (0) | 0% (0) | 0.051% (3) | 0.04% (1) |
| Progressive supranuclear ophthalmoplegia [Steele-Richardson-Olszewski] | 0.005% (1) | 0% (0) | 0% (0) | 0.023% (1) | 0% (0) | 0% (0) |
| Psoriasis and psoriatic arthritis | 0.197% (42) | 0.396% (13) | 0.336% (18) | 0.117% (5) | 0.068% (4) | 0.08% (2) |
| Psychotic disorder with hallucinations due to known physiological condition | 0.005% (1) | 0.03% (1) | 0% (0) | 0% (0) | 0% (0) | 0% (0) |
| Pulmonary Fibrosis | 0.239% (51) | 0.396% (13) | 0.261% (14) | 0.21% (9) | 0.187% (11) | 0.16% (4) |
| Reactive attachment disorder of childhood | 0.005% (1) | 0% (0) | 0.019% (1) | 0% (0) | 0% (0) | 0% (0) |
| Restless legs syndrome | 0.258% (55) | 0.122% (4) | 0.261% (14) | 0.327% (14) | 0.306% (18) | 0.2% (5) |
| Rheumatoid Arthritis | 0.737% (157) | 0.761% (25) | 0.821% (44) | 0.63% (27) | 0.732% (43) | 0.719% (18) |
| Schizoaffective disorder, unspecified | 0.366% (78) | 0.396% (13) | 0.485% (26) | 0.21% (9) | 0.425% (25) | 0.2% (5) |
| Schizoaffective disorders | 0.455% (97) | 0.457% (15) | 0.634% (34) | 0.303% (13) | 0.459% (27) | 0.32% (8) |
| Schizophrenia | 0.77% (164) | 0.974% (32) | 0.914% (49) | 0.513% (22) | 0.715% (42) | 0.759% (19) |
| Sepsis | 6.808% (1451) | 10.35% (340) | 7.966% (427) | 8.141% (349) | 4.067% (239) | 3.835% (96) |
| Shock, unspecified | 0.727% (155) | 1.37% (45) | 0.914% (49) | 0.84% (36) | 0.34% (20) | 0.2% (5) |
| Shortness of breath | 18.285% (3897) | 22.374% (735) | 23.993% (1286) | 24.283% (1041) | 10.277% (604) | 9.229% (231) |
| Social phobia, unspecified | 0.014% (3) | 0% (0) | 0% (0) | 0% (0) | 0.051% (3) | 0% (0) |
| Social phobias | 0.014% (3) | 0% (0) | 0% (0) | 0% (0) | 0.051% (3) | 0% (0) |
| Specific (isolated) phobias | 0.014% (3) | 0% (0) | 0.037% (2) | 0% (0) | 0% (0) | 0.04% (1) |
| Spinal muscular atrophy and related syndromes | 0.009% (2) | 0% (0) | 0% (0) | 0% (0) | 0.017% (1) | 0.04% (1) |
| Tachycardia, unspecified | 3.979% (848) | 4.627% (152) | 5.429% (291) | 4.735% (203) | 2.382% (140) | 2.477% (62) |
| Thrombocytopenia, unspecified | 2.684% (572) | 3.653% (120) | 2.929% (157) | 3.242% (139) | 1.974% (116) | 1.598% (40) |
| Tic disorder | 0.009% (2) | 0% (0) | 0.019% (1) | 0% (0) | 0.017% (1) | 0% (0) |
| Tourette's disorder | 0.009% (2) | 0% (0) | 0.019% (1) | 0% (0) | 0.017% (1) | 0% (0) |
| Trigeminal neuralgia | 0.08% (17) | 0.061% (2) | 0.112% (6) | 0.07% (3) | 0.085% (5) | 0.04% (1) |
| Tuberous sclerosis | 0.005% (1) | 0% (0) | 0% (0) | 0% (0) | 0.017% (1) | 0% (0) |
| Type 2 diabetes | 18.074% (3852) | 28.463% (935) | 22.836% (1224) | 16.912% (725) | 12.813% (753) | 8.59% (215) |
| Unspecified atrial fibrillation | 3.796% (809) | 3.531% (116) | 4.16% (223) | 4.105% (176) | 3.42% (201) | 3.716% (93) |
| Unspecified dementia | 2.196% (468) | 3.105% (102) | 3.097% (166) | 1.213% (52) | 1.736% (102) | 1.838% (46) |
| Unspecified disorder of psychological development | 0.005% (1) | 0.03% (1) | 0% (0) | 0% (0) | 0% (0) | 0% (0) |
| Unspecified disorder of vestibular function | 0.005% (1) | 0% (0) | 0% (0) | 0% (0) | 0.017% (1) | 0% (0) |
| Unspecified protein-calorie malnutrition | 0.577% (123) | 0.639% (21) | 0.616% (33) | 0.606% (26) | 0.562% (33) | 0.4% (10) |
| Unspecified severe protein-calorie malnutrition | 1.093% (233) | 1.461% (48) | 1.082% (58) | 0.933% (40) | 0.987% (58) | 1.159% (29) |
| UTI | 3.299% (703) | 3.957% (130) | 3.507% (188) | 2.286% (98) | 3.471% (204) | 3.316% (83) |
| Vitamin B12 deficiency anemia due to intrinsic factor deficiency | 0.005% (1) | 0.03% (1) | 0% (0) | 0% (0) | 0% (0) | 0% (0) |
| Vitamin D deficiency, unspecified | 0.615% (131) | 0.944% (31) | 1.007% (54) | 0.373% (16) | 0.391% (23) | 0.28% (7) |
| Volume depletion | 7.334% (1563) | 9.802% (322) | 8.601% (461) | 7.068% (303) | 5.275% (310) | 6.672% (167) |
| Wernicke's encephalopathy | 0.009% (2) | 0% (0) | 0.019% (1) | 0.023% (1) | 0% (0) | 0% (0) |

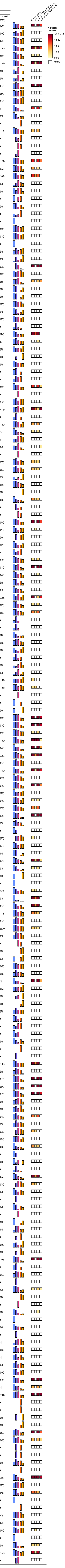

### Table S3 - Consultations by COVID-19 Wave

| Consultations | Total | Alpha/Beta | | Delta | Omicron | |
|---|---|---|---|---|---|---|
| | | Wave 1 (03-01-2020 to 10-01-2020) | Wave 2 (10-01-2020 to 06-15-2021) | Wave 3 (06-15-2021 to 11-15-2021) | Wave 4 (11-15-2021 to 04-01-2022) | Wave 5 (04-01-2022 to 09-01-2022) |
| Anesthesia | 0.774% (165) | 0.913% (30) | 0.746% (40) | 0.77% (33) | 0.8% (47) | 0.599% (15) |
| Cardiology | 4.533% (966) | 5.388% (177) | 4.459% (239) | 3.522% (151) | 4.696% (276) | 4.914% (123) |
| Critical Care/Pulmonary | 5.171% (1102) | 8.98% (295) | 5.765% (309) | 6.368% (273) | 2.859% (168) | 2.277% (57) |
| Endocrine | 0.239% (51) | 0.304% (10) | 0.299% (16) | 0.187% (8) | 0.272% (16) | 0.04% (1) |
| Gastroenterology | 2.933% (625) | 3.623% (119) | 3.246% (174) | 2.613% (112) | 2.671% (157) | 2.517% (63) |
| Hem-Onc | 1.056% (225) | 1.705% (56) | 0.951% (51) | 0.933% (40) | 0.987% (58) | 0.799% (20) |
| Infectious Disease | 3.496% (745) | 6.332% (208) | 3.899% (209) | 3.196% (137) | 2.552% (150) | 1.638% (41) |
| Nephrology | 4.2% (895) | 6.454% (212) | 4.776% (256) | 3.965% (170) | 3.114% (183) | 2.956% (74) |
| Neurology | 3.669% (782) | 4.262% (140) | 4.403% (236) | 2.729% (117) | 3.709% (218) | 2.837% (71) |
| Neurosurgery | 0.962% (205) | 1.187% (39) | 0.989% (53) | 0.956% (41) | 0.902% (53) | 0.759% (19) |
| Orthopedic | 1.29% (275) | 1.735% (57) | 1.493% (80) | 1.026% (44) | 1.361% (80) | 0.559% (14) |
| Palliative Care | 5.87% (1251) | 10.046% (330) | 6.698% (359) | 6.718% (288) | 3.505% (206) | 2.717% (68) |
| Physical Medicine and Rehab | 0.882% (188) | 1.035% (34) | 0.746% (40) | 0.816% (35) | 0.919% (54) | 0.999% (25) |
| Psychiatry | 2.036% (434) | 2.435% (80) | 2.388% (128) | 1.563% (67) | 1.753% (103) | 2.237% (56) |
| Rheumatology | 0.178% (38) | 0.365% (12) | 0.187% (10) | 0.14% (6) | 0.17% (10) | 0% (0) |
| Surgery | 1.028% (219) | 1.431% (47) | 1.063% (57) | 0.933% (40) | 1.004% (59) | 0.639% (16) |
| Urology | 0.61% (130) | 0.852% (28) | 0.634% (34) | 0.467% (20) | 0.596% (35) | 0.519% (13) |

Frequency

Global p.value
Wave 1-2 vs Wave 3
Wave 1-2 vs Wave 4-5
Wave 3 vs Wave 4-5

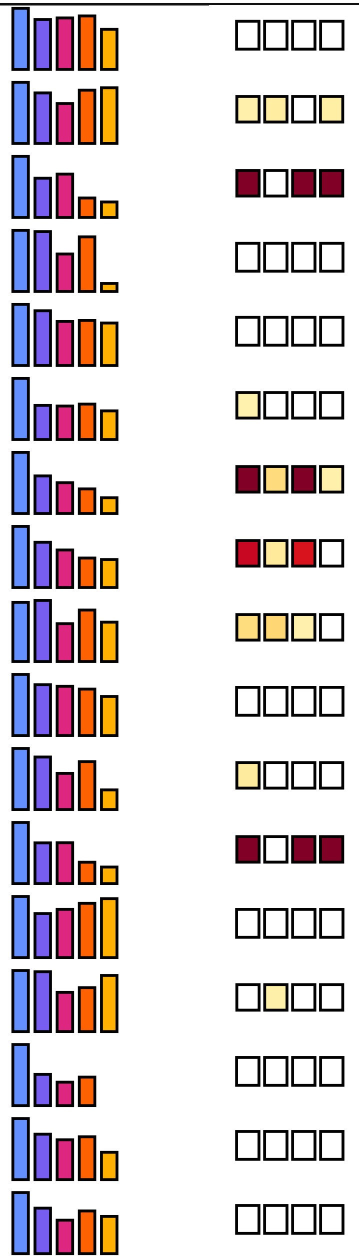

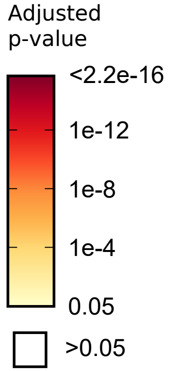

### Table S4 - Diagnoses by Trajectory Group

| Diagnoses | Total (21312) | Moderate TG1A (17476) | Severe TG1B (1351) | Severe TG1C (701) | Critical TG2 (715) | Critical TG3 (380) | Death TG4 (689) |
|---|---|---|---|---|---|---|---|
| Acidosis | 5.607% (1195) | 2.369% (414) | 12.879% (174) | 14.551% (102) | 20% (143) | 29.474% (112) | 36.284% (250) |
| Actively Intoxicated | 0.511% (109) | 0.532% (93) | 0.592% (8) | 0.428% (3) | 0.42% (3) | 0% (0) | 0.29% (2) |
| Acute and chronic respiratory failure with hypoxia | 1.206% (257) | 0.309% (54) | 3.849% (52) | 2.996% (21) | 5.874% (42) | 10.526% (40) | 6.967% (48) |
| Acute kidney failure | 10.098% (2152) | 4.572% (799) | 21.688% (293) | 30.1% (211) | 33.287% (238) | 54.737% (208) | 58.491% (403) |
| Acute posthemorrhagic anemia | 1.412% (301) | 0.515% (90) | 2.591% (35) | 4.708% (33) | 6.853% (49) | 17.105% (65) | 4.209% (29) |
| Acute respiratory distress | 19.576% (4172) | 7.948% (1389) | 64.619% (873) | 58.06% (407) | 78.741% (563) | 90.526% (344) | 86.502% (596) |
| Adjustment disorder with depressed mood | 0.042% (9) | 0.017% (3) | 0.074% (1) | 0.285% (2) | 0.14% (1) | 0.263% (1) | 0.145% (1) |
| Adjustment disorders | 0.131% (28) | 0.069% (12) | 0.37% (5) | 0.571% (4) | 0.28% (2) | 1.053% (4) | 0.145% (1) |
| Adverse effect of glucocorticoids and synthetic analogues, initial encounter | 3.735% (796) | 0.927% (162) | 13.768% (186) | 14.551% (102) | 21.399% (153) | 26.842% (102) | 13.208% (91) |
| Alcohol Abuse | 1.905% (406) | 1.625% (284) | 3.553% (48) | 3.138% (22) | 4.476% (32) | 1.579% (6) | 2.032% (14) |
| Alcohol related disorders | 1.905% (406) | 1.625% (284) | 3.553% (48) | 3.138% (22) | 4.476% (32) | 1.579% (6) | 2.032% (14) |
| Alkalosis | 0.981% (209) | 0.183% (32) | 1.628% (22) | 2.282% (16) | 5.594% (40) | 18.158% (69) | 4.354% (30) |
| Alzheimer's disease | 0.338% (72) | 0.217% (38) | 0.444% (6) | 1.141% (8) | 0.839% (6) | 0% (0) | 2.032% (14) |
| Amnestic disorder due to known physiological condition | 0.005% (1) | 0.006% (1) | 0% (0) | 0% (0) | 0% (0) | 0% (0) | 0% (0) |
| Anemia | 5.856% (1248) | 3.531% (617) | 10.881% (147) | 14.123% (99) | 18.462% (132) | 33.947% (129) | 17.997% (124) |
| Ankylosing spondylitis | 0.028% (6) | 0.017% (3) | 0.148% (2) | 0% (0) | 0.14% (1) | 0% (0) | 0% (0) |
| Anorexia nervosa | 0.009% (2) | 0.006% (1) | 0% (0) | 0.143% (1) | 0% (0) | 0% (0) | 0% (0) |
| Antisocial personality disorder | 0.014% (3) | 0.011% (2) | 0% (0) | 0.143% (1) | 0% (0) | 0% (0) | 0% (0) |
| Anxiety disorder, unspecified | 6.691% (1426) | 4.063% (710) | 13.62% (184) | 16.405% (115) | 21.119% (151) | 33.421% (127) | 20.174% (139) |
| Asthma | 5.335% (1137) | 4.881% (853) | 8.29% (112) | 7.418% (52) | 7.692% (55) | 7.105% (27) | 5.515% (38) |
| Athereosclretic heart disease | 5.861% (1249) | 3.725% (651) | 11.917% (161) | 15.977% (112) | 16.084% (115) | 12.105% (46) | 23.803% (164) |
| Attention-deficit hyperactivity disorders | 0.343% (73) | 0.32% (56) | 0.592% (8) | 0.571% (4) | 0.559% (4) | 0% (0) | 0.145% (1) |
| Autistic disorder | 0.075% (16) | 0.057% (10) | 0.148% (2) | 0.285% (2) | 0% (0) | 0% (0) | 0.29% (2) |
| Autoimmune Hepatitis | 0.033% (7) | 0.029% (5) | 0.148% (2) | 0% (0) | 0% (0) | 0% (0) | 0% (0) |
| Bell's palsy | 0.089% (19) | 0.063% (11) | 0.074% (1) | 0.285% (2) | 0.42% (3) | 0.263% (1) | 0.145% (1) |
| Benign intracranial hypertension | 0.033% (7) | 0.017% (3) | 0% (0) | 0% (0) | 0% (0) | 0.263% (1) | 0.435% (3) |
| Binge eating disorder | 0.005% (1) | 0.006% (1) | 0% (0) | 0% (0) | 0% (0) | 0% (0) | 0% (0) |
| Bipolar | 1.68% (358) | 1.373% (240) | 3.035% (41) | 4.422% (31) | 2.937% (21) | 2.632% (10) | 2.177% (15) |
| Bipolar disorder | 1.68% (358) | 1.373% (240) | 3.035% (41) | 4.422% (31) | 2.937% (21) | 2.632% (10) | 2.177% (15) |
| Borderline intellectual functioning | 0.005% (1) | 0% (0) | 0% (0) | 0.143% (1) | 0% (0) | 0% (0) | 0% (0) |
| Borderline personality disorder | 0.122% (26) | 0.092% (16) | 0.148% (2) | 0.713% (5) | 0.42% (3) | 0% (0) | 0% (0) |
| Brief psychotic disorder | 0.122% (26) | 0.074% (13) | 0.37% (5) | 0.856% (6) | 0.28% (2) | 0% (0) | 0% (0) |
| Burn-out | 4.833% (1030) | 2.804% (490) | 15.914% (215) | 12.696% (89) | 13.566% (97) | 13.158% (50) | 12.917% (89) |
| COPD | 3.12% (665) | 1.843% (322) | 8.142% (110) | 7.989% (56) | 9.231% (66) | 5.526% (21) | 13.062% (90) |
| Cannabis related disorders | 0.859% (183) | 0.687% (120) | 1.925% (26) | 2.282% (16) | 1.399% (10) | 1.842% (7) | 0.581% (4) |
| Cardiac arrest, cause unspecified | 0.995% (212) | 0.017% (3) | 0.222% (3) | 0.143% (1) | 1.818% (13) | 10% (38) | 22.351% (154) |
| Celiac Disease | 0.033% (7) | 0.034% (6) | 0% (0) | 0% (0) | 0% (0) | 0.263% (1) | 0% (0) |
| Celiac disease | 0.033% (7) | 0.034% (6) | 0% (0) | 0% (0) | 0% (0) | 0.263% (1) | 0% (0) |
| Cerebral infarction | 0.957% (204) | 0.355% (62) | 2.221% (30) | 4.422% (31) | 3.636% (26) | 6.316% (24) | 4.499% (31) |
| Cerebral palsy | 0.141% (30) | 0.063% (11) | 0.74% (10) | 0.285% (2) | 0.559% (4) | 0.526% (2) | 0.145% (1) |
| Cerebral palsy and other paralytic syndromes | 0.948% (202) | 0.286% (50) | 2.073% (28) | 3.709% (26) | 4.615% (33) | 10.263% (39) | 3.774% (26) |
| Chronic Kidney Disease | 3.918% (835) | 2.255% (394) | 10.437% (141) | 10.556% (74) | 10.769% (77) | 8.684% (33) | 16.836% (116) |
| Chronic inflammatory demyelinating polyneuritis | 0.033% (7) | 0% (0) | 0% (0) | 0.143% (1) | 0.559% (4) | 0.263% (1) | 0.145% (1) |
| Chronic obstructive pulmonary disease with (acute) lower respiratory infection | 1.567% (334) | 0.561% (98) | 5.107% (69) | 4.137% (29) | 6.853% (49) | 5.263% (20) | 10.015% (69) |
| Chronic pain syndrome | 0.216% (46) | 0.109% (19) | 0.592% (8) | 0.571% (4) | 1.119% (8) | 0.789% (3) | 0.581% (4) |
| Cluster headaches and other trigeminal autonomic cephalgias (TAC) | 0.009% (2) | 0.006% (1) | 0.074% (1) | 0% (0) | 0% (0) | 0% (0) | 0% (0) |
| Cocaine related disorders | 0.469% (100) | 0.343% (60) | 0.814% (11) | 1.712% (12) | 0.979% (7) | 1.579% (6) | 0.581% (4) |
| Conduct disorder, unspecified | 0.019% (4) | 0.011% (2) | 0% (0) | 0% (0) | 0.28% (2) | 0% (0) | 0% (0) |
| Conduct disorders | 0.042% (9) | 0.034% (6) | 0% (0) | 0% (0) | 0.42% (3) | 0% (0) | 0% (0) |
| Constipation | 3.308% (705) | 1.076% (188) | 8.216% (111) | 14.551% (102) | 20.979% (150) | 21.316% (81) | 10.595% (73) |
| Conversion disorder with motor symptom or deficit | 0.005% (1) | 0.006% (1) | 0% (0) | 0% (0) | 0% (0) | 0% (0) | 0% (0) |
| Cough | 16.094% (3430) | 17.55% (3067) | 11.251% (152) | 8.987% (63) | 10.629% (76) | 9.474% (36) | 5.225% (36) |
| Degenerative disease of nervous system, unspecified | 0.033% (7) | 0.017% (3) | 0.222% (3) | 0.143% (1) | 0% (0) | 0% (0) | 0% (0) |
| Dehydration | 5.748% (1225) | 4.309% (753) | 10.955% (148) | 13.267% (93) | 14.965% (107) | 10.789% (41) | 12.046% (83) |
| Delirium due to known physiological condition | 0.521% (111) | 0.103% (18) | 1.036% (14) | 2.853% (20) | 3.077% (22) | 6.053% (23) | 2.032% (14) |
| Delusional disorders | 0.183% (39) | 0.126% (22) | 0.444% (6) | 0.856% (6) | 0.28% (2) | 0.263% (1) | 0.29% (2) |
| Dependence on respirator [ventilator] status | 0.338% (72) | 0.011% (2) | 0.074% (1) | 0.143% (1) | 0.699% (5) | 13.947% (53) | 1.451% (10) |
| Dependent personality disorder | 0.005% (1) | 0.006% (1) | 0% (0) | 0% (0) | 0% (0) | 0% (0) | 0% (0) |
| Depression | 4.978% (1061) | 3.759% (657) | 9.623% (130) | 12.268% (86) | 10.769% (77) | 12.895% (49) | 8.999% (62) |
| Depressive episode | 4.978% (1061) | 3.759% (657) | 9.623% (130) | 12.268% (86) | 10.769% (77) | 12.895% (49) | 8.999% (62) |
| Disappearance and death of family member | 0.08% (17) | 0.04% (7) | 0.222% (3) | 0.143% (1) | 0.28% (2) | 0.526% (2) | 0.29% (2) |
| Disorders of calcium metabolism | 0.967% (206) | 0.446% (78) | 2.443% (33) | 2.425% (17) | 4.336% (31) | 5% (19) | 4.064% (28) |
| Disorders of copper metabolism | 0.005% (1) | 0% (0) | 0% (0) | 0% (0) | 0.14% (1) | 0% (0) | 0% (0) |
| Disorders of phosphorus metabolism and phosphatases | 1.154% (246) | 0.383% (67) | 2.517% (34) | 3.566% (25) | 3.916% (28) | 10.526% (40) | 7.547% (52) |
| Dissociative and conversion disorder, unspecified | 0.033% (7) | 0.023% (4) | 0.074% (1) | 0% (0) | 0.28% (2) | 0% (0) | 0% (0) |
| Dissociative identity disorder | 0.009% (2) | 0.011% (2) | 0% (0) | 0% (0) | 0% (0) | 0% (0) | 0% (0) |
| Do not resuscitate | 6.203% (1322) | 1.579% (276) | 10.659% (144) | 13.695% (96) | 19.021% (136) | 34.211% (130) | 78.374% (540) |
| Dysphagia, unspecified | 1.553% (331) | 0.486% (85) | 2.443% (33) | 5.278% (37) | 10.49% (75) | 15% (57) | 6.386% (44) |
| Dysthymic disorder | 0.023% (5) | 0.023% (4) | 0.074% (1) | 0% (0) | 0% (0) | 0% (0) | 0% (0) |
| Elevated blood-pressure | 0.572% (122) | 0.612% (107) | 0.518% (7) | 0.143% (1) | 0.699% (5) | 0.263% (1) | 0.145% (1) |
| Encephalitis, myelitis and encephalomyelitis, unspecified | 0.047% (10) | 0.017% (3) | 0.074% (1) | 0.285% (2) | 0.14% (1) | 0.526% (2) | 0.145% (1) |
| Encephalopathy | 3.205% (683) | 1.07% (187) | 6.514% (88) | 10.984% (77) | 15.944% (114) | 24.474% (93) | 17.997% (124) |
| Encounter for palliative care | 4.57% (974) | 0.343% (60) | 3.849% (52) | 7.418% (52) | 15.944% (114) | 48.158% (183) | 74.456% (513) |
| Epilepsy and recurrent seizures | 1.45% (309) | 0.944% (165) | 3.331% (45) | 4.28% (30) | 4.056% (29) | 4.737% (18) | 3.193% (22) |
| Facial nerve disorders | 0.089% (19) | 0.063% (11) | 0.074% (1) | 0.285% (2) | 0.42% (3) | 0.263% (1) | 0.145% (1) |
| Fatty (change of) liver, not elsewhere classified | 0.746% (159) | 0.463% (81) | 2.073% (28) | 2.14% (15) | 1.538% (11) | 3.158% (12) | 1.742% (12) |
| Fever, unspecified | 9.896% (2109) | 10.117% (1768) | 8.734% (118) | 10.699% (75) | 8.531% (61) | 11.579% (44) | 6.241% (43) |
| Fluid overload, unspecified | 1.178% (251) | 0.326% (57) | 1.925% (26) | 3.424% (24) | 5.175% (37) | 12.368% (47) | 8.708% (60) |
| Gastro-esophageal reflux disease without esophagitis | 4.481% (955) | 3.021% (528) | 9.697% (131) | 9.7% (68) | 13.846% (99) | 12.368% (47) | 11.901% (82) |
| Gender identity disorder, unspecified | 0.028% (6) | 0.023% (4) | 0% (0) | 0.143% (1) | 0% (0) | 0.263% (1) | 0% (0) |
| Gender identity disorders | 0.028% (6) | 0.023% (4) | 0% (0) | 0.143% (1) | 0% (0) | 0.263% (1) | 0% (0) |
| Generalized anxiety disorder | 0.385% (82) | 0.257% (45) | 0.592% (8) | 0.999% (7) | 1.259% (9) | 1.579% (6) | 1.016% (7) |
| Gout | 0.882% (188) | 0.475% (83) | 2.221% (30) | 2.425% (17) | 2.797% (20) | 3.684% (14) | 3.483% (24) |
| Grave's Disease | 0.047% (10) | 0.051% (9) | 0.074% (1) | 0% (0) | 0% (0) | 0% (0) | 0% (0) |
| Guillain-Barre syndrome | 0.033% (7) | 0.017% (3) | 0% (0) | 0.143% (1) | 0.28% (2) | 0.263% (1) | 0% (0) |
| Hallucinogen related disorders | 0.047% (10) | 0.046% (8) | 0.074% (1) | 0% (0) | 0.14% (1) | 0% (0) | 0% (0) |
| Hashimoto's Throiditis | 0.108% (23) | 0.092% (16) | 0.296% (4) | 0% (0) | 0.28% (2) | 0.263% (1) | 0% (0) |
| Headache | 4.096% (873) | 4.583% (801) | 2.147% (29) | 1.712% (12) | 2.937% (21) | 1.053% (4) | 0.871% (6) |
| Heart Failure | 5.748% (1225) | 3.216% (562) | 13.768% (186) | 15.977% (112) | 17.203% (123) | 19.211% (73) | 24.528% (169) |
| Hereditary and idiopathic neuropathy, unspecified | 0.005% (1) | 0.006% (1) | 0% (0) | 0% (0) | 0% (0) | 0% (0) | 0% (0) |
| Hereditary motor and sensory neuropathy | 0.009% (2) | 0.011% (2) | 0% (0) | 0% (0) | 0% (0) | 0% (0) | 0% (0) |
| Huntington's disease | 0.009% (2) | 0.006% (1) | 0% (0) | 0% (0) | 0% (0) | 0% (0) | 0.145% (1) |
| Hyperglycemia | 4.049% (863) | 2.386% (417) | 11.029% (149) | 10.699% (75) | 11.189% (80) | 17.895% (68) | 10.74% (74) |
| Hyperglycemia, unspecified | 4.049% (863) | 2.386% (417) | 11.029% (149) | 10.699% (75) | 11.189% (80) | 17.895% (68) | 10.74% (74) |
| Hyperkalemia | 3.463% (738) | 1.144% (200) | 5.699% (77) | 10.128% (71) | 12.727% (91) | 27.632% (105) | 28.157% (194) |
| Hyperlipidemia, unspecified | 12.383% (2639) | 7.731% (1351) | 29.978% (405) | 31.526% (221) | 33.986% (243) | 31.316% (119) | 43.541% (300) |
| Hyperosmolality and hypernatremia | 2.501% (533) | 0.326% (57) | 2.073% (28) | 4.565% (32) | 13.007% (93) | 45% (171) | 22.061% (152) |
| Hypertension | 21.978% (4684) | 17.035% (2977) | 40.785% (551) | 43.367% (304) | 47.133% (337) | 51.053% (194) | 46.589% (321) |
| Hypertensive heart disease with heart failure | 2.801% (597) | 1.677% (293) | 6.884% (93) | 8.274% (58) | 7.692% (55) | 7.632% (29) | 10.015% (69) |
| Hypo-osmolality and hyponatremia | 7.306% (1557) | 3.376% (590) | 19.689% (266) | 22.967% (161) | 29.51% (211) | 44.737% (170) | 23.077% (159) |
| Hypoglycemia, unspecified | 0.755% (161) | 0.315% (55) | 1.554% (21) | 2.425% (17) | 3.776% (27) | 5.526% (21) | 2.903% (20) |
| Hypokalemia | 5.809% (1238) | 3.491% (610) | 14.804% (200) | 14.123% (99) | 20.28% (145) | 25.526% (97) | 12.627% (87) |
| Hypomagnesemia | 1.014% (216) | 0.578% (101) | 2.887% (39) | 3.138% (22) | 3.077% (22) | 4.474% (17) | 2.177% (15) |
| Hypotension | 1.811% (386) | 0.795% (139) | 3.183% (43) | 5.278% (37) | 7.273% (52) | 12.368% (47) | 9.869% (68) |
| Hypothyroidism | 4.819% (1027) | 3.33% (582) | 11.029% (149) | 10.556% (74) | 11.608% (83) | 8.947% (34) | 15.239% (105) |
| Hypoxemia | 9.516% (2028) | 5.104% (892) | 30.2% (408) | 28.673% (201) | 30.35% (217) | 27.368% (104) | 29.898% (206) |
| Ileus, unspecified | 0.432% (92) | 0.109% (19) | 0.444% (6) | 1.141% (8) | 1.678% (12) | 10.263% (39) | 1.161% (8) |
| Impulse disorder, unspecified | 0.009% (2) | 0.011% (2) | 0% (0) | 0% (0) | 0% (0) | 0% (0) | 0% (0) |
| Imunnodeficiency | 0.629% (134) | 0.326% (57) | 1.554% (21) | 1.284% (9) | 2.098% (15) | 4.211% (16) | 2.322% (16) |
| Inflammatory Bowel Disease | 0.948% (202) | 0.692% (121) | 1.85% (25) | 2.14% (15) | 2.657% (19) | 2.632% (10) | 1.742% (12) |
| Insomnia not due to a substance or known physiological condition | 0.047% (10) | 0.017% (3) | 0.074% (1) | 0.428% (3) | 0.42% (3) | 0% (0) | 0% (0) |
| Insulin Use | 6.658% (1419) | 3.988% (697) | 16.284% (220) | 18.83% (132) | 22.517% (161) | 18.421% (70) | 20.174% (139) |
| Intellectual disabilities | 0.286% (61) | 0.172% (30) | 0.74% (10) | 1.284% (9) | 1.119% (8) | 0.263% (1) | 0.435% (3) |
| Intermittent explosive disorder | 0.009% (2) | 0.011% (2) | 0% (0) | 0% (0) | 0% (0) | 0% (0) | 0% (0) |
| Intracranial abscess and granuloma | 0.005% (1) | 0% (0) | 0% (0) | 0% (0) | 0% (0) | 0.263% (1) | 0% (0) |
| Iron deficiency anemia, unspecified | 1.633% (348) | 1.076% (188) | 4.071% (55) | 2.996% (21) | 6.154% (44) | 4.737% (18) | 3.193% (22) |
| Liver disease | 1.136% (242) | 0.538% (94) | 3.553% (48) | 3.566% (25) | 4.895% (35) | 5.789% (22) | 2.612% (18) |
| Long Term Coagulants | 3.96% (844) | 2.518% (440) | 9.474% (128) | 9.843% (69) | 12.727% (91) | 10.526% (40) | 11.03% (76) |
| Long term (current) use of oral hypoglycemic drugs | 4.974% (1060) | 4.126% (721) | 8.66% (117) | 10.556% (74) | 9.371% (67) | 6.842% (26) | 7.983% (55) |
| Long term asprin | 6.405% (1365) | 4.646% (812) | 12.139% (164) | 19.544% (137) | 14.406% (103) | 8.421% (32) | 16.981% (117) |
| Long term drug therapy | 17.624% (3756) | 15.175% (2652) | 29.238% (395) | 33.951% (238) | 29.93% (214) | 28.421% (108) | 21.626% (149) |
| Lupus | 0.347% (74) | 0.252% (44) | 0.592% (8) | 0.856% (6) | 1.399% (10) | 0.526% (2) | 0.581% (4) |
| Magnesium deficiency | 0.009% (2) | 0.011% (2) | 0% (0) | 0% (0) | 0% (0) | 0% (0) | 0% (0) |
| Malignant neoplasm of brain, unspecified | 0.014% (3) | 0% (0) | 0% (0) | 0.285% (2) | 0.14% (1) | 0% (0) | 0% (0) |
| Malingerer [conscious simulation] | 0.033% (7) | 0.029% (5) | 0.074% (1) | 0.143% (1) | 0% (0) | 0% (0) | 0% (0) |
| Malnutrition | 2.102% (448) | 0.652% (114) | 4.959% (67) | 7.703% (54) | 11.608% (83) | 18.947% (72) | 8.418% (58) |
| Migraine | 0.99% (211) | 0.83% (145) | 1.554% (21) | 1.854% (13) | 2.797% (20) | 1.579% (6) | 0.871% (6) |
| Mixed disorder of acid-base balance | 1.248% (266) | 0.103% (18) | 1.184% (16) | 1.712% (12) | 4.056% (29) | 29.211% (111) | 11.611% (80) |
| Moderate protein-calorie malnutrition | 0.371% (79) | 0.097% (17) | 1.48% (20) | 1.427% (10) | 1.399% (10) | 3.158% (12) | 1.451% (10) |
| Mood disorder due to known physiological condition | 0.014% (3) | 0.006% (1) | 0.074% (1) | 0.143% (1) | 0% (0) | 0% (0) | 0% (0) |
| Motor neuron disease | 0.009% (2) | 0.011% (2) | 0% (0) | 0% (0) | 0% (0) | 0% (0) | 0% (0) |
| Multiple Sclerosis | 0.136% (29) | 0.12% (21) | 0.37% (5) | 0.285% (2) | 0.14% (1) | 0% (0) | 0% (0) |
| Myasthenia gravis | 0.038% (8) | 0.011% (2) | 0.148% (2) | 0.143% (1) | 0.14% (1) | 0% (0) | 0.29% (2) |
| Myoclonus | 0.103% (22) | 0.023% (4) | 0.074% (1) | 0.713% (5) | 0.14% (1) | 0.789% (3) | 1.161% (8) |
| Narcolepsy and cataplexy | 0.014% (3) | 0.011% (2) | 0% (0) | 0% (0) | 0% (0) | 0.263% (1) | 0% (0) |
| Neurocognitive disorder with Lewy bodies | 0.056% (12) | 0.017% (3) | 0.296% (4) | 0.143% (1) | 0.28% (2) | 0% (0) | 0.29% (2) |
| Neuromyelitis optica [Devic] | 0.009% (2) | 0.006% (1) | 0% (0) | 0.143% (1) | 0% (0) | 0% (0) | 0% (0) |
| Niacin deficiency [pellagra] | 0.005% (1) | 0.006% (1) | 0% (0) | 0% (0) | 0% (0) | 0% (0) | 0% (0) |
| Nictoine Dependence | 6.808% (1451) | 4.349% (760) | 17.691% (239) | 17.261% (121) | 18.182% (130) | 11.316% (43) | 22.932% (158) |
| Nontraumatic intracerebral hemorrhage | 0.249% (53) | 0.04% (7) | 0.148% (2) | 1.141% (8) | 0.839% (6) | 2.105% (8) | 3.193% (22) |
| Obesity | 8.418% (1794) | 5.11% (893) | 19.763% (267) | 19.116% (134) | 27.413% (196) | 37.368% (142) | 23.512% (162) |
| Obesity due to excess calories | 3.5% (746) | 1.728% (302) | 8.586% (116) | 10.842% (76) | 13.007% (93) | 22.105% (84) | 10.885% (75) |
| Obesity, unspecified | 5.213% (1111) | 3.491% (610) | 11.695% (158) | 8.845% (62) | 15.944% (114) | 17.895% (68) | 14.369% (99) |
| Obsessive-compulsive disorder | 0.075% (16) | 0.051% (9) | 0.148% (2) | 0% (0) | 0.559% (4) | 0.263% (1) | 0% (0) |
| Obsessive-compulsive disorder, unspecified | 0.066% (14) | 0.046% (8) | 0.148% (2) | 0% (0) | 0.42% (3) | 0.263% (1) | 0% (0) |
| Obstructive sleep apnea | 2.599% (554) | 1.253% (219) | 7.254% (98) | 7.133% (50) | 11.329% (81) | 10.526% (40) | 9.579% (66) |
| Opioid related disorders | 0.38% (81) | 0.315% (55) | 0.444% (6) | 1.284% (9) | 0.839% (6) | 0.526% (2) | 0.435% (3) |
| Other abnormalities of plasma proteins | 0.957% (204) | 0.629% (110) | 1.628% (22) | 1.427% (10) | 3.636% (26) | 2.895% (11) | 3.628% (25) |
| Other disorders of electrolyte and fluid balance, not elsewhere classified | 0.704% (150) | 0.372% (65) | 2.665% (36) | 1.854% (13) | 2.098% (15) | 3.158% (12) | 1.306% (9) |
| Other disorders of phosphorus metabolism | 1.154% (246) | 0.383% (67) | 2.517% (34) | 3.566% (25) | 3.916% (28) | 10.526% (40) | 7.547% (52) |
| Other eating disorders | 0.009% (2) | 0.006% (1) | 0.074% (1) | 0% (0) | 0% (0) | 0% (0) | 0% (0) |
| Other encephalitis and encephalomyelitis | 0.028% (6) | 0.011% (2) | 0% (0) | 0.285% (2) | 0% (0) | 0.526% (2) | 0% (0) |
| Other inflammatory polyneuropathies | 0.038% (8) | 0.006% (1) | 0% (0) | 0.143% (1) | 0.559% (4) | 0.263% (1) | 0.145% (1) |
| Other specified abnormal findings of blood chemistry | 2.886% (615) | 1.911% (334) | 6.588% (89) | 6.562% (46) | 6.853% (49) | 7.368% (28) | 10.015% (69) |
| Other specified abnormalities of plasma proteins | 0.957% (204) | 0.629% (110) | 1.628% (22) | 1.427% (10) | 3.636% (26) | 2.895% (11) | 3.628% (25) |
| Other specified anxiety disorders | 0.108% (23) | 0.092% (16) | 0.222% (3) | 0% (0) | 0.42% (3) | 0.263% (1) | 0% (0) |
| Other specified eating disorder | 0.005% (1) | 0% (0) | 0.074% (1) | 0% (0) | 0% (0) | 0% (0) | 0% (0) |
| Other specified metabolic disorders | 0.122% (26) | 0.069% (12) | 0.148% (2) | 0.713% (5) | 0.699% (5) | 0.263% (1) | 0.145% (1) |
| Other sphingolipidosis | 0.005% (1) | 0.006% (1) | 0% (0) | 0% (0) | 0% (0) | 0% (0) | 0% (0) |
| Other vitamin deficiencies | 0.019% (4) | 0.006% (1) | 0.074% (1) | 0.143% (1) | 0% (0) | 0.263% (1) | 0% (0) |
| Panic disorder [episodic paroxysmal anxiety] | 0.399% (85) | 0.229% (40) | 0.814% (11) | 1.141% (8) | 1.119% (8) | 3.158% (12) | 0.871% (6) |
| Paranoid personality disorder | 0.014% (3) | 0.011% (2) | 0% (0) | 0.143% (1) | 0% (0) | 0% (0) | 0% (0) |
| Parkinson's disease | 0.436% (93) | 0.2% (35) | 1.036% (14) | 1.569% (11) | 1.538% (11) | 0.263% (1) | 3.048% (21) |
| Phobic anxiety disorders | 0.033% (7) | 0.029% (5) | 0.074% (1) | 0% (0) | 0.14% (1) | 0% (0) | 0% (0) |
| Pneumonia | 29.087% (6199) | 17.973% (3141) | 74.537% (1007) | 67.618% (474) | 82.937% (593) | 95.526% (363) | 90.131% (621) |
| Polyarteritis Nodosa | 0.019% (4) | 0.011% (2) | 0.074% (1) | 0.143% (1) | 0% (0) | 0% (0) | 0% (0) |
| Post-traumatic stress disorder (PTSD) | 0.784% (167) | 0.635% (111) | 1.776% (24) | 1.427% (10) | 0.979% (7) | 2.368% (9) | 0.871% (6) |
| Postsurgical malabsorption, not elsewhere classified | 0.005% (1) | 0% (0) | 0% (0) | 0% (0) | 0% (0) | 0% (0) | 0.145% (1) |
| Prediabetes | 0.652% (139) | 0.401% (70) | 1.702% (23) | 1.284% (9) | 2.378% (17) | 2.105% (8) | 1.742% (12) |
| Primary Biliary Cirrhosis | 0.019% (4) | 0.017% (3) | 0.074% (1) | 0% (0) | 0% (0) | 0% (0) | 0% (0) |
| Progressive supranuclear ophthalmoplegia [Steele-Richardson-Olszewski] | 0.005% (1) | 0.006% (1) | 0% (0) | 0% (0) | 0% (0) | 0% (0) | 0% (0) |
| Psoriasis and psoriatic arthritis | 0.197% (42) | 0.137% (24) | 0.37% (5) | 0.713% (5) | 0.559% (4) | 0.263% (1) | 0.435% (3) |
| Psychotic disorder with hallucinations due to known physiological condition | 0.005% (1) | 0% (0) | 0% (0) | 0% (0) | 0% (0) | 0.263% (1) | 0% (0) |
| Pulmonary Fibrosis | 0.239% (51) | 0.074% (13) | 0.222% (3) | 0.571% (4) | 0.839% (6) | 3.158% (12) | 1.887% (13) |
| Reactive attachment disorder of childhood | 0.005% (1) | 0% (0) | 0% (0) | 0.143% (1) | 0% (0) | 0% (0) | 0% (0) |
| Restless legs syndrome | 0.258% (55) | 0.114% (20) | 1.036% (14) | 1.141% (8) | 0.42% (3) | 0.526% (2) | 1.161% (8) |
| Rheumatoid Arthritis | 0.737% (157) | 0.504% (88) | 1.258% (17) | 1.712% (12) | 1.958% (14) | 1.579% (6) | 2.903% (20) |
| Schizoaffective disorder, unspecified | 0.366% (78) | 0.252% (44) | 1.036% (14) | 0.999% (7) | 0.979% (7) | 0.789% (3) | 0.435% (3) |
| Schizoaffective disorders | 0.455% (97) | 0.298% (52) | 1.258% (17) | 1.569% (11) | 1.399% (10) | 0.789% (3) | 0.581% (4) |
| Schizophrenia | 0.77% (164) | 0.618% (108) | 1.702% (23) | 1.854% (13) | 1.818% (13) | 0.526% (2) | 0.726% (5) |
| Sepsis | 6.808% (1451) | 2.466% (431) | 15.396% (208) | 19.544% (137) | 28.252% (202) | 53.421% (203) | 39.187% (270) |
| Shock, unspecified | 0.727% (155) | 0.046% (8) | 0.518% (7) | 1.141% (8) | 2.378% (17) | 10.789% (41) | 10.74% (74) |
| Shortness of breath | 18.285% (3897) | 14.477% (2530) | 37.38% (505) | 32.525% (228) | 34.266% (245) | 33.947% (129) | 37.736% (260) |
| Social phobia, unspecified | 0.014% (3) | 0.006% (1) | 0.074% (1) | 0% (0) | 0.14% (1) | 0% (0) | 0% (0) |
| Social phobias | 0.014% (3) | 0.006% (1) | 0.074% (1) | 0% (0) | 0.14% (1) | 0% (0) | 0% (0) |
| Specific (isolated) phobias | 0.014% (3) | 0.017% (3) | 0% (0) | 0% (0) | 0% (0) | 0% (0) | 0% (0) |
| Spinal muscular atrophy and related syndromes | 0.009% (2) | 0.011% (2) | 0% (0) | 0% (0) | 0% (0) | 0% (0) | 0% (0) |
| Tachycardia, unspecified | 3.979% (848) | 3.439% (601) | 5.699% (77) | 6.419% (45) | 7.552% (54) | 10.526% (40) | 4.499% (31) |
| Thrombocytopenia, unspecified | 2.684% (572) | 1.316% (230) | 4.959% (67) | 6.134% (43) | 8.671% (62) | 19.737% (75) | 13.788% (95) |
| Tic disorder | 0.009% (2) | 0.006% (1) | 0% (0) | 0% (0) | 0% (0) | 0% (0) | 0.145% (1) |
| Tourette's disorder | 0.009% (2) | 0.006% (1) | 0% (0) | 0% (0) | 0% (0) | 0% (0) | 0.145% (1) |
| Trigeminal neuralgia | 0.08% (17) | 0.069% (12) | 0.148% (2) | 0.143% (1) | 0.14% (1) | 0% (0) | 0.145% (1) |
| Tuberous sclerosis | 0.005% (1) | 0.006% (1) | 0% (0) | 0% (0) | 0% (0) | 0% (0) | 0% (0) |
| Type 2 diabetes | 18.074% (3852) | 12.589% (2200) | 37.454% (506) | 38.802% (272) | 45.874% (328) | 53.158% (202) | 49.927% (344) |
| UTI | 3.299% (703) | 2.42% (423) | 5.625% (76) | 6.277% (44) | 8.951% (64) | 8.158% (31) | 9.434% (65) |
| Unspecified atrial fibrillation | 3.796% (809) | 2.06% (360) | 6.44% (87) | 8.845% (62) | 13.706% (98) | 11.579% (44) | 22.932% (158) |
| Unspecified dementia | 2.196% (468) | 1.087% (190) | 5.774% (78) | 7.846% (55) | 5.734% (41) | 4.474% (17) | 12.627% (87) |
| Unspecified disorder of psychological development | 0.005% (1) | 0% (0) | 0% (0) | 0.143% (1) | 0% (0) | 0% (0) | 0% (0) |
| Unspecified disorder of vestibular function | 0.005% (1) | 0.006% (1) | 0% (0) | 0% (0) | 0% (0) | 0% (0) | 0% (0) |
| Unspecified protein-calorie malnutrition | 0.577% (123) | 0.177% (31) | 1.11% (15) | 2.14% (15) | 3.357% (24) | 5.789% (22) | 2.322% (16) |
| Unspecified severe protein-calorie malnutrition | 1.093% (233) | 0.309% (54) | 2.147% (29) | 4.565% (32) | 6.853% (49) | 10% (38) | 4.499% (31) |
| Vitamin B12 deficiency anemia due to intrinsic factor deficiency | 0.005% (1) | 0.006% (1) | 0% (0) | 0% (0) | 0% (0) | 0% (0) | 0% (0) |
| Vitamin D deficiency, unspecified | 0.615% (131) | 0.349% (61) | 1.85% (25) | 2.425% (17) | 1.958% (14) | 1.842% (7) | 1.016% (7) |
| Volume depletion | 7.334% (1563) | 4.99% (872) | 14.286% (193) | 18.973% (133) | 22.238% (159) | 21.316% (81) | 18.142% (125) |
| Wernicke's encephalopathy | 0.009% (2) | 0% (0) | 0% (0) | 0.285% (2) | 0% (0) | 0% (0) | 0% (0) |

Adjusted p-value: <2.2e-16, 1e-12, 1e-8, 1e-4, 0.05; >0.05

### Table S5 - Consultations by Trajectory Group

| Consultations | Total (21312) | Moderate TG1A (17476) | Severe TG1B (1351) | Severe TG1C (701) | Critical TG2 (715) | Critical TG3 (380) | Death TG4 (689) | Frequency | Distribution | Global p.value / TG1A vs TG1B-4 / TG1A vs TG1B-1C / TG3 vs TG4 / TG2 vs TG3 |
|---|---|---|---|---|---|---|---|---|---|---|
| Anesthesia | 0.774% (165) | 0.658% (115) | 1.184% (16) | 1.997% (14) | 1.678% (12) | 0.789% (3) | 0.726% (5) | | | |
| Cardiology | 4.533% (966) | 2.735% (478) | 8.808% (119) | 11.983% (84) | 15.385% (110) | 20% (76) | 14.369% (99) | | | |
| Critical Care/Pulmonary | 5.171% (1102) | 0.973% (170) | 6.884% (93) | 13.409% (94) | 32.028% (229) | 59.474% (226) | 42.09% (290) | | | |
| Endocrine | 0.239% (51) | 0.166% (29) | 0.592% (8) | 0.285% (2) | 0.979% (7) | 0.526% (2) | 0.435% (3) | | | |
| Gastroenterology | 2.933% (625) | 1.654% (289) | 5.551% (75) | 7.846% (55) | 11.189% (80) | 25.263% (96) | 4.354% (30) | | | |
| Hem-Onc | 1.056% (225) | 0.555% (97) | 1.702% (23) | 3.566% (25) | 4.336% (31) | 8.947% (34) | 2.177% (15) | | | |
| Infectious Disease | 3.496% (745) | 1.333% (233) | 7.106% (96) | 11.27% (79) | 14.266% (102) | 36.316% (138) | 14.078% (97) | | | |
| Nephrology | 4.2% (895) | 1.751% (306) | 7.476% (101) | 10.128% (71) | 12.168% (87) | 31.842% (121) | 30.334% (209) | | | |
| Neurology | 3.669% (782) | 2.22% (388) | 6.218% (84) | 10.699% (75) | 11.469% (82) | 20.526% (78) | 10.885% (75) | | | |
| Neurosurgery | 0.962% (205) | 0.578% (101) | 1.776% (24) | 3.424% (24) | 3.077% (22) | 3.421% (13) | 3.048% (21) | | | |
| Orthopedic | 1.29% (275) | 0.956% (167) | 3.257% (44) | 3.709% (26) | 3.497% (25) | 1.316% (5) | 1.161% (8) | | | |
| Palliative Care | 5.87% (1251) | 1.081% (189) | 5.922% (80) | 12.981% (91) | 24.476% (175) | 71.316% (271) | 64.586% (445) | | | |
| Physical Medicine and Rehab | 0.882% (188) | 0.446% (78) | 1.85% (25) | 2.853% (20) | 4.336% (31) | 8.684% (33) | 0.145% (1) | | | |
| Psychiatry | 2.036% (434) | 1.339% (234) | 4.589% (62) | 7.275% (51) | 5.315% (38) | 10.526% (40) | 1.306% (9) | | | |
| Rheumatology | 0.178% (38) | 0.132% (23) | 0.222% (3) | 0.713% (5) | 0.699% (5) | 0.263% (1) | 0.145% (1) | | | |
| Surgery | 1.028% (219) | 0.71% (124) | 1.332% (18) | 3.281% (23) | 3.497% (25) | 5% (19) | 1.451% (10) | | | |
| Urology | 0.61% (130) | 0.326% (57) | 1.554% (21) | 2.71% (19) | 2.797% (20) | 2.632% (10) | 0.435% (3) | | | |

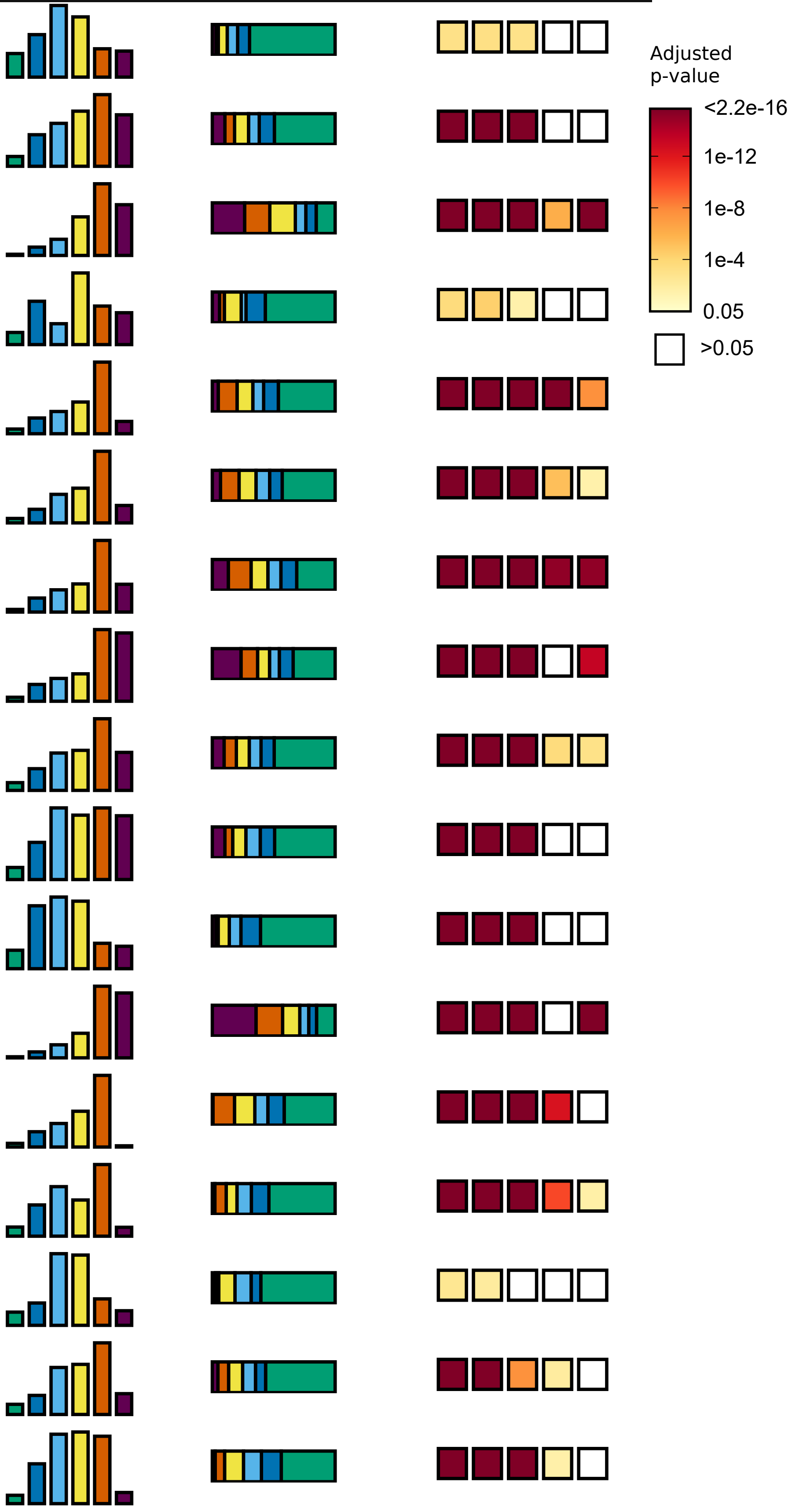

### Table S6 - Imaging Studies by Trajectory Group

| Imaging Study | Total (21312) | Moderate TG1A (17476) | Severe TG1B (1351) | Severe TG1C (701) | Critical TG2 (715) | Critical TG3 (380) | Death TG4 (689) |
|---|---|---|---|---|---|---|---|
| CT Angiography Chest | 12.007% (2559) | 8.549% (1494) | 22.428% (303) | 25.392% (178) | 34.685% (248) | 47.632% (181) | 22.496% (155) |
| CT Chest w/o Contrast | 2.454% (523) | 1.493% (261) | 4.885% (66) | 5.849% (41) | 6.434% (46) | 16.842% (64) | 6.531% (45) |
| CT Head w/o Contrast | 10.684% (2277) | 7.656% (1338) | 17.691% (239) | 25.535% (179) | 26.154% (187) | 38.684% (147) | 27.141% (187) |
| US LE Venous Duplex Bilat | 4.035% (860) | 1.539% (269) | 6.44% (87) | 9.986% (70) | 19.161% (137) | 43.684% (166) | 19.013% (131) |
| US RENAL | 2.487% (530) | 1.265% (221) | 5.774% (78) | 7.989% (56) | 8.392% (60) | 8.684% (33) | 11.901% (82) |
| XR Abdomen AP | 6.935% (1478) | 2.106% (368) | 7.846% (106) | 14.979% (105) | 32.727% (234) | 83.684% (318) | 50.363% (347) |
| XR Chest 1 View | 57.381% (12229) | 50.292% (8789) | 81.939% (1107) | 88.445% (620) | 94.825% (678) | 99.474% (378) | 95.356% (657) |
| XR Speech Evaluation Dyname Pharyngeal | 1.811% (386) | 0.584% (102) | 2.517% (34) | 5.706% (40) | 11.748% (84) | 28.158% (107) | 2.758% (19) |

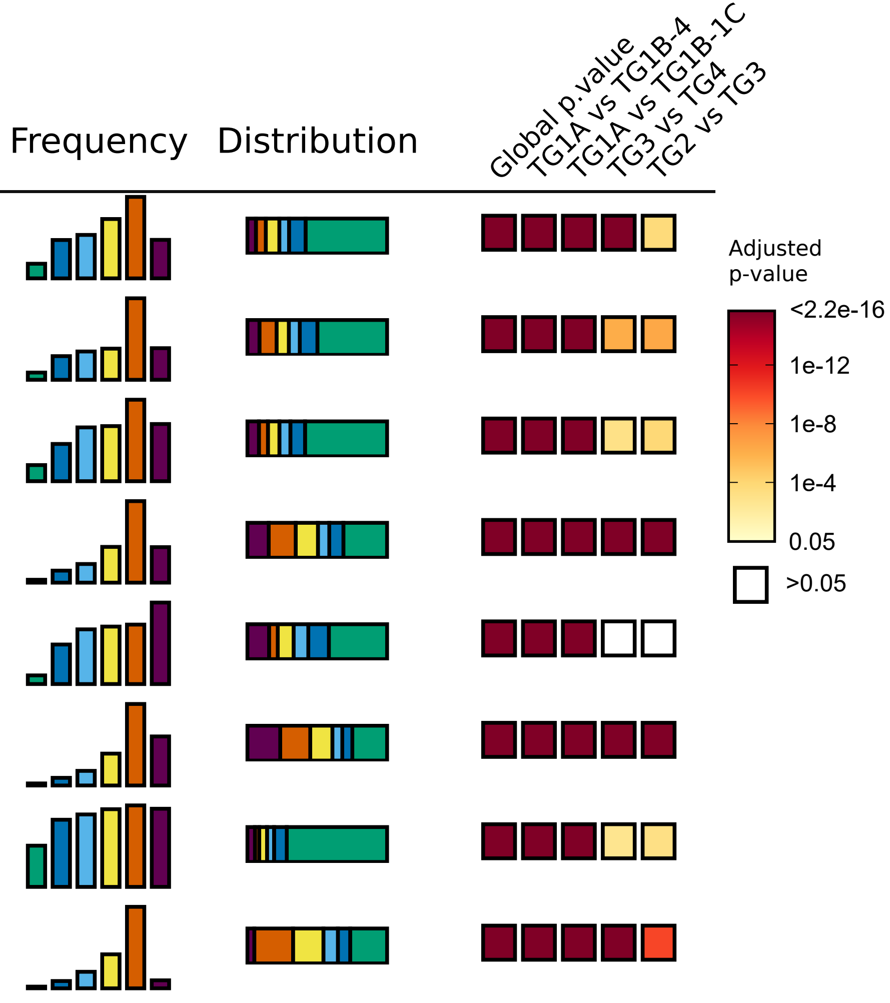

Table S7 - Administered Medications by Trajectory Group

| Administered Medications | Total (21312) | Moderate TG1A (17476) | Severe TG1B (1351) | Severe TG1C (701) | Critical TG2 (715) | Critical TG3 (380) | Death TG4 (689) |
|---|---|---|---|---|---|---|---|
| acetaminophen | 54.692% (11656) | 45.663% (7980) | 95.633% (1292) | 96.291% (675) | 97.902% (700) | 97.895% (372) | 92.453% (637) |
| acetaminophen-HYDROcodone | 6.288% (1340) | 4.303% (752) | 13.397% (181) | 14.836% (104) | 19.58% (140) | 18.684% (71) | 13.353% (92) |
| acetaZOLAMIDE | 0.845% (180) | 0.057% (10) | 0.444% (6) | 1.569% (11) | 2.937% (21) | 25.789% (98) | 4.935% (34) |
| aerochamber | 6.846% (1459) | 3.468% (606) | 20.947% (283) | 18.26% (128) | 24.615% (176) | 29.211% (111) | 22.496% (155) |
| Al hydroxide/Mg hydroxide/simethicone | 21.448% (4571) | 15.255% (2666) | 52.702% (712) | 54.066% (379) | 48.531% (347) | 42.105% (160) | 44.557% (307) |
| albumin human | 1.882% (401) | 0.223% (39) | 1.554% (21) | 4.28% (30) | 9.091% (65) | 31.316% (119) | 18.433% (127) |
| albuterol | 20.467% (4362) | 12.451% (2176) | 56.847% (768) | 49.501% (347) | 59.161% (423) | 64.737% (246) | 58.345% (402) |
| albuterol-ipratropium | 2.721% (580) | 0.841% (147) | 5.107% (69) | 7.133% (50) | 14.685% (105) | 26.316% (100) | 15.82% (109) |
| ALPRAZolam | 1.717% (366) | 0.675% (118) | 4.441% (60) | 5.136% (36) | 8.112% (58) | 13.947% (53) | 5.951% (41) |
| alteplase | 19.89% (4239) | 12.182% (2129) | 48.409% (654) | 54.636% (383) | 60.559% (433) | 67.632% (257) | 55.588% (383) |
| AMIOdarone | 1.605% (342) | 0.355% (62) | 1.554% (21) | 3.709% (26) | 6.154% (44) | 14.211% (54) | 19.594% (135) |
| amLODIPine | 7.142% (1522) | 4.189% (732) | 18.505% (250) | 21.255% (149) | 23.077% (165) | 26.579% (101) | 18.142% (125) |
| ampicillin-sulbactam | 1.469% (313) | 0.566% (99) | 1.85% (25) | 5.421% (38) | 8.252% (59) | 14.211% (54) | 5.515% (38) |
| apixaban | 3.824% (815) | 2.089% (365) | 11.103% (150) | 10.984% (77) | 18.322% (131) | 10% (38) | 7.837% (54) |
| aspirin | 11.017% (2348) | 7.696% (1345) | 23.316% (315) | 27.104% (190) | 28.811% (206) | 23.947% (91) | 29.173% (201) |
| atorvastatin | 11.501% (2451) | 6.724% (1175) | 30.57% (413) | 30.956% (217) | 35.804% (256) | 38.158% (145) | 35.559% (245) |
| atracurium | 2.773% (591) | 0.006% (1) | 0.296% (4) | 0.856% (6) | 11.329% (81) | 62.632% (238) | 37.881% (261) |
| atropine | 1.492% (318) | 0.28% (49) | 1.332% (18) | 2.14% (15) | 5.734% (41) | 18.421% (70) | 18.142% (125) |
| azithromycin | 12.481% (2660) | 7.382% (1290) | 31.458% (425) | 32.382% (227) | 43.776% (313) | 48.421% (184) | 32.075% (221) |
| balsam Peru-castor oil topical | 1.882% (401) | 0.28% (49) | 2.369% (32) | 4.85% (34) | 7.133% (51) | 29.211% (111) | 17.997% (124) |
| benzonatate | 26.924% (5738) | 19.072% (3333) | 64.101% (866) | 61.341% (430) | 63.636% (455) | 68.421% (260) | 57.184% (394) |
| bisacodyl | 27.252% (5808) | 18.208% (3182) | 61.362% (829) | 67.475% (473) | 69.79% (499) | 87.368% (332) | 71.553% (493) |
| calcium | 4.082% (870) | 1.476% (258) | 7.328% (99) | 11.412% (80) | 14.545% (104) | 30% (114) | 31.205% (215) |
| carvedilol | 3.163% (674) | 1.928% (337) | 7.476% (101) | 10.414% (73) | 9.371% (67) | 7.895% (30) | 9.579% (66) |
| ceFAZolin | 7.719% (1645) | 5.499% (961) | 11.103% (150) | 16.976% (119) | 20.839% (149) | 39.211% (149) | 16.981% (117) |
| cefepime | 6.076% (1295) | 1.282% (224) | 8.364% (113) | 14.123% (99) | 30.35% (217) | 77.368% (294) | 50.508% (348) |
| cefTRIAXone | 13.232% (2820) | 7.09% (1239) | 34.641% (468) | 37.66% (264) | 46.993% (336) | 58.947% (224) | 41.945% (289) |
| cholecalciferol | 3.669% (782) | 1.865% (326) | 11.769% (159) | 11.412% (80) | 15.804% (113) | 11.579% (44) | 8.708% (60) |
| cholecalciferol (Vitamin D3 (Cholecalciferol)) | 2.984% (636) | 1.442% (252) | 9.919% (134) | 9.986% (70) | 13.846% (99) | 8.158% (31) | 7.257% (50) |
| codeine-guaiFENesin | 4.167% (888) | 2.272% (397) | 12.435% (168) | 11.27% (79) | 13.986% (100) | 17.895% (68) | 11.03% (76) |
| dexamethasone | 27.975% (5962) | 16.978% (2967) | 74.463% (1006) | 68.616% (481) | 84.056% (601) | 88.947% (338) | 82.583% (569) |
| dexmedetomidine | 2.853% (608) | 0.2% (35) | 2.073% (28) | 5.136% (36) | 18.741% (134) | 56.316% (214) | 23.367% (161) |
| dextromethorphan-guaiFENesin | 10.032% (2138) | 5.951% (1040) | 28.793% (389) | 25.82% (181) | 32.587% (233) | 30.789% (117) | 25.835% (178) |
| dextrose | 6.076% (1295) | 1.465% (256) | 7.994% (108) | 14.836% (104) | 26.993% (193) | 66.842% (254) | 55.152% (380) |
| diphenhydrAMINE | 11.477% (2446) | 9.55% (1669) | 21.392% (289) | 23.538% (165) | 24.615% (176) | 18.684% (71) | 11.03% (76) |
| docusate | 8.521% (1816) | 5.848% (1022) | 11.769% (159) | 18.26% (128) | 23.916% (171) | 43.684% (166) | 24.673% (170) |
| docusate-senna | 21.631% (4610) | 13.624% (2381) | 53.812% (727) | 58.631% (411) | 59.301% (424) | 67.895% (258) | 59.361% (409) |
| emollients, topical | 3.472% (740) | 3.256% (569) | 2.443% (33) | 4.137% (29) | 3.217% (23) | 15% (57) | 4.209% (29) |
| enoxaparin | 27.74% (5912) | 16.743% (2926) | 78.608% (1062) | 77.318% (542) | 83.077% (594) | 88.421% (336) | 65.602% (452) |
| EPINEPHrine | 2.703% (576) | 0.83% (145) | 2.369% (32) | 4.708% (33) | 9.231% (66) | 23.684% (90) | 30.479% (210) |
| EPINEPHrine-lidocaine | 1.858% (396) | 1.305% (228) | 1.925% (26) | 5.136% (36) | 6.294% (45) | 11.316% (43) | 2.612% (18) |
| etomidate | 1.539% (328) | 0.092% (16) | 0.814% (11) | 2.282% (16) | 8.531% (61) | 30.789% (117) | 15.53% (107) |
| famotidine | 12.223% (2605) | 5.991% (1047) | 24.796% (335) | 29.672% (208) | 44.615% (319) | 78.684% (299) | 57.62% (397) |
| fentaNYL | 13.645% (2908) | 8.68% (1517) | 13.916% (188) | 25.25% (177) | 41.818% (299) | 84.211% (320) | 59.071% (407) |
| fluticasone | 2.309% (492) | 1.116% (195) | 6.81% (92) | 7.989% (56) | 11.189% (80) | 6.316% (24) | 6.531% (45) |
| furosemide | 11.214% (2390) | 3.096% (541) | 26.573% (359) | 36.234% (254) | 61.958% (443) | 90.526% (344) | 65.167% (449) |
| gabapentin | 5.659% (1206) | 3.611% (631) | 13.101% (177) | 17.546% (123) | 20.28% (145) | 14.211% (54) | 11.03% (76) |
| glucagon | 2.482% (529) | 0.652% (114) | 3.997% (54) | 8.417% (59) | 12.727% (91) | 26.053% (99) | 16.255% (112) |
| glycopyrrolate | 2.872% (612) | 1.35% (236) | 2.813% (38) | 4.422% (31) | 8.112% (58) | 8.158% (31) | 31.64% (218) |
| guaiFENesin | 18.145% (3867) | 10.552% (1844) | 51.739% (699) | 50.214% (352) | 58.881% (421) | 59.211% (225) | 47.315% (326) |
| haloperidol | 2.252% (480) | 0.893% (156) | 4.071% (55) | 5.706% (40) | 9.65% (69) | 9.211% (35) | 18.142% (125) |
| heparin | 10.196% (2173) | 4.858% (849) | 22.354% (302) | 29.672% (208) | 37.483% (268) | 53.158% (202) | 49.927% (344) |
| hydrALAZINE | 8.723% (1859) | 4.732% (827) | 20.873% (282) | 23.68% (166) | 30.49% (218) | 40% (152) | 31.06% (214) |
| hydrocortisone | 0.906% (193) | 0.223% (39) | 0.74% (10) | 0.999% (7) | 2.797% (20) | 10.789% (41) | 11.03% (76) |
| HYDROmorphone | 8.31% (1771) | 6.34% (1108) | 12.879% (174) | 20.685% (145) | 22.937% (164) | 21.842% (83) | 14.078% (97) |
| hydrOXYzine | 4.95% (1055) | 2.718% (475) | 12.953% (175) | 16.262% (114) | 20% (143) | 17.368% (66) | 11.901% (82) |
| ibuprofen | 14.091% (3003) | 14.202% (2482) | 15.544% (210) | 17.261% (121) | 14.126% (101) | 13.684% (52) | 5.37% (37) |
| influenza virus vaccine, inactivated | 3.575% (762) | 2.409% (421) | 9.104% (123) | 9.986% (70) | 10.49% (75) | 11.053% (42) | 4.499% (31) |
| insulin | 20.078% (4279) | 11.627% (2032) | 46.336% (626) | 51.07% (358) | 65.315% (467) | 84.474% (321) | 68.94% (475) |
| iohexol | 12.007% (2559) | 8.555% (1495) | 21.466% (290) | 26.534% (186) | 34.825% (249) | 44.474% (169) | 24.673% (170) |
| ipratropium | 1.609% (343) | 0.813% (142) | 2.739% (37) | 3.566% (25) | 7.413% (53) | 10.789% (41) | 6.531% (45) |
| IV Fluids | 52.585% (11207) | 43.328% (7572) | 90.896% (1228) | 93.866% (658) | 97.063% (694) | 100% (380) | 97.968% (675) |
| labetalol | 3.402% (725) | 1.762% (308) | 6.218% (84) | 11.127% (78) | 10.909% (78) | 22.632% (86) | 13.208% (91) |
| lactulose | 1.778% (379) | 0.452% (79) | 2.813% (38) | 5.563% (39) | 11.049% (79) | 19.474% (74) | 10.16% (70) |
| levothyroxine | 4.209% (897) | 2.541% (444) | 11.991% (162) | 10.414% (73) | 11.329% (81) | 8.947% (34) | 14.949% (103) |
| lidocaine | 19.951% (4252) | 12.034% (2103) | 30.496% (412) | 50.785% (356) | 72.587% (519) | 94.474% (359) | 73.004% (503) |
| lisinopril | 5.171% (1102) | 3.193% (558) | 13.472% (182) | 16.405% (115) | 14.965% (107) | 17.105% (65) | 10.885% (75) |
| loperamide | 5.56% (1185) | 2.781% (486) | 16.062% (217) | 20.399% (143) | 20% (143) | 19.211% (73) | 17.852% (123) |
| LORazepam | 9.253% (1972) | 4.721% (825) | 15.766% (213) | 21.541% (151) | 26.294% (188) | 42.105% (160) | 63.135% (435) |
| losartan | 3.083% (657) | 1.86% (325) | 8.882% (120) | 7.703% (54) | 10.769% (77) | 7.895% (30) | 7.402% (51) |
| magnesium | 22.269% (4746) | 12.938% (2261) | 54.774% (740) | 61.056% (428) | 71.748% (513) | 86.579% (329) | 68.94% (475) |
| melatonin | 10.051% (2142) | 4.767% (833) | 27.683% (374) | 35.521% (249) | 42.937% (307) | 44.211% (168) | 30.624% (211) |
| metFORMIN | 2.825% (602) | 1.711% (299) | 8.216% (111) | 8.417% (59) | 11.608% (83) | 8.158% (31) | 2.758% (19) |
| methylPREDNISolone | 2.473% (527) | 0.916% (160) | 6.07% (82) | 5.421% (38) | 11.888% (85) | 22.632% (86) | 11.03% (76) |
| metoCLOPRAMIDE | 2.079% (443) | 1.373% (240) | 2.147% (29) | 2.853% (20) | 6.713% (48) | 17.105% (65) | 5.951% (41) |
| metoPROLOL | 6.963% (1484) | 3.491% (610) | 16.136% (218) | 18.26% (128) | 25.175% (180) | 40.263% (153) | 28.302% (195) |
| metroNIDAZOLE | 2.022% (431) | 1.207% (211) | 2.517% (34) | 6.134% (43) | 6.154% (44) | 11.842% (45) | 7.837% (54) |
| midazolam | 8.577% (1828) | 4.126% (721) | 8.512% (115) | 16.405% (115) | 32.308% (231) | 79.211% (301) | 50.073% (345) |
| morphine | 12.139% (2587) | 8.658% (1513) | 21.095% (285) | 19.401% (136) | 24.196% (173) | 27.368% (104) | 54.572% (376) |
| multivitamin | 6.771% (1443) | 4.709% (823) | 15.766% (213) | 17.689% (124) | 18.601% (133) | 20.263% (77) | 10.595% (73) |
| mupirocin topical | 2.548% (543) | 0.767% (134) | 6.292% (85) | 8.417% (59) | 12.587% (90) | 17.105% (65) | 15.965% (110) |
| nitric oxide | 0.788% (168) | 0% (0) | 0.074% (1) | 0.285% (2) | 1.538% (11) | 19.474% (74) | 11.611% (80) |
| norepinephrine | 4.692% (1000) | 0.332% (58) | 1.999% (27) | 5.849% (41) | 23.916% (171) | 76.579% (291) | 59.797% (412) |
| ocular lubricant | 4.35% (927) | 0.532% (93) | 3.627% (49) | 5.706% (40) | 21.678% (155) | 69.474% (264) | 47.315% (326) |
| OLANZapine | 2.698% (575) | 1.036% (181) | 5.403% (73) | 9.7% (68) | 11.888% (85) | 26.842% (102) | 9.579% (66) |
| ondansetron | 45.763% (9753) | 36.513% (6381) | 89.563% (1210) | 90.585% (635) | 88.392% (632) | 86.579% (329) | 82.148% (566) |
| oxyCODONE | 11.242% (2396) | 8.703% (1521) | 20.947% (283) | 27.532% (193) | 29.371% (210) | 28.158% (107) | 11.901% (82) |
| oxymetazoline nasal | 0.854% (182) | 0.257% (45) | 1.184% (16) | 1.569% (11) | 4.895% (35) | 10.789% (41) | 4.935% (34) |
| pancrelipase | 4.58% (976) | 0.223% (39) | 2.369% (32) | 7.846% (55) | 28.392% (203) | 77.895% (296) | 50.943% (351) |
| pantoprazole | 9.736% (2075) | 5.27% (921) | 25.833% (349) | 26.534% (186) | 33.566% (240) | 36.842% (140) | 34.688% (239) |
| perflutren | 2.876% (613) | 1.167% (204) | 4.145% (56) | 8.559% (60) | 14.126% (101) | 24.474% (93) | 14.369% (99) |
| phenylephrine | 7.423% (1582) | 4.223% (738) | 6.218% (84) | 12.411% (87) | 21.119% (151) | 58.947% (224) | 43.251% (298) |
| piperacillin-tazobactam | 3.519% (750) | 1.476% (258) | 6.144% (83) | 9.986% (70) | 14.965% (107) | 25% (95) | 19.884% (137) |
| polyethylene glycol 3350 | 8.845% (1885) | 2.689% (470) | 19.097% (258) | 29.957% (210) | 48.531% (347) | 83.158% (316) | 41.219% (284) |
| Potassium | 24.897% (5306) | 14.677% (2565) | 62.398% (843) | 66.191% (464) | 79.441% (568) | 92.632% (352) | 74.601% (514) |
| predniSONE | 3.097% (660) | 2.06% (360) | 6.44% (87) | 6.705% (47) | 11.469% (82) | 13.947% (53) | 4.499% (31) |
| PrismaSol dialysis solution | 0.582% (124) | 0.011% (2) | 0% (0) | 0.143% (1) | 1.119% (8) | 11.316% (43) | 10.16% (70) |
| promethazine | 18.694% (3984) | 15.038% (2628) | 34.789% (470) | 39.515% (277) | 37.622% (269) | 35.789% (136) | 29.608% (204) |
| propofol | 9.117% (1943) | 4.452% (778) | 9.919% (134) | 18.83% (132) | 35.105% (251) | 80.526% (306) | 49.637% (342) |
| QUEtiapine | 1.52% (324) | 0.595% (104) | 3.405% (46) | 4.708% (33) | 9.371% (67) | 13.421% (51) | 3.338% (23) |
| remdesivir | 12.364% (2635) | 4.4% (769) | 47.372% (640) | 40.514% (284) | 51.189% (366) | 61.842% (235) | 49.492% (341) |
| rocuronium | 5.537% (1180) | 2.392% (418) | 5.329% (72) | 9.843% (69) | 23.217% (166) | 62.105% (236) | 31.785% (219) |
| scopolamine | 0.971% (207) | 0.235% (41) | 0.666% (9) | 0.999% (7) | 2.937% (21) | 2.895% (11) | 17.126% (118) |
| senna | 2.679% (571) | 0.847% (148) | 6.218% (84) | 10.128% (71) | 17.063% (122) | 23.158% (88) | 8.418% (58) |
| sodium bicarbonate | 7.691% (1639) | 4.011% (701) | 7.032% (95) | 13.98% (98) | 23.357% (167) | 56.579% (215) | 52.685% (363) |
| sodium chrloride nebs | 2.567% (547) | 0.36% (63) | 3.849% (52) | 5.706% (40) | 18.601% (133) | 39.737% (151) | 15.675% (108) |
| sodium phosphate | 12.908% (2751) | 6.523% (1140) | 32.05% (433) | 35.663% (250) | 46.853% (335) | 65.263% (248) | 50.073% (345) |
| sodium polystyrene sulfonate | 1.403% (299) | 0.298% (52) | 2.073% (28) | 2.71% (19) | 5.315% (38) | 14.737% (56) | 15.385% (106) |
| succinylcholine | 2.416% (515) | 1.007% (176) | 2.517% (34) | 4.565% (32) | 11.329% (81) | 27.632% (105) | 12.627% (87) |
| tamsulosin | 3.205% (683) | 1.762% (308) | 7.032% (95) | 11.698% (82) | 12.168% (87) | 13.158% (50) | 8.853% (61) |
| tocilizumab | 2.065% (440) | 0.092% (16) | 5.181% (70) | 6.562% (46) | 15.105% (108) | 21.053% (80) | 17.417% (120) |
| traMADol | 4.645% (990) | 3.079% (538) | 10.659% (144) | 13.124% (92) | 13.566% (97) | 12.368% (47) | 10.45% (72) |
| traZODone | 2.853% (608) | 1.465% (256) | 6.958% (94) | 8.559% (60) | 13.986% (100) | 10.526% (40) | 8.418% (58) |
| vancomycin | 6.123% (1305) | 1.791% (313) | 9.623% (130) | 17.261% (121) | 28.252% (202) | 64.737% (246) | 42.525% (293) |
| vasopressin | 2.313% (493) | 0.16% (28) | 1.258% (17) | 1.427% (10) | 6.434% (46) | 32.368% (123) | 39.042% (269) |
| vecuronium | 0.68% (145) | 0.011% (2) | 0.518% (7) | 0.999% (7) | 2.517% (18) | 18.421% (70) | 5.951% (41) |
| zinc | 2.473% (527) | 0.921% (161) | 6.958% (94) | 7.275% (51) | 12.587% (90) | 15.263% (58) | 10.595% (73) |

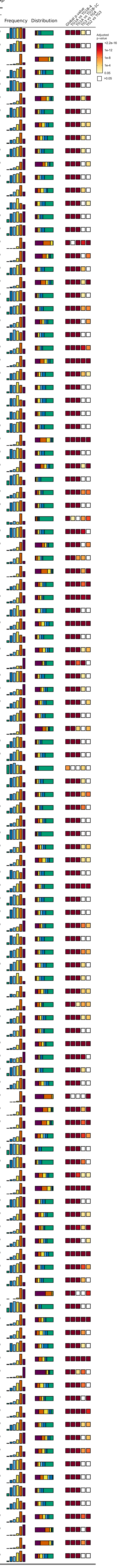

### Table S8 - Labratory Tests by Trajectory Group

| Panel | Laboratory Test | Total (21312) | Moderate TG1A (17476) | Severe TG1B (1351) | Severe TG1C (701) | Critical TG2 (715) | Critical TG3 (380) | Death TG4 (689) |
|---|---|---|---|---|---|---|---|---|
| Activated PTT | PTT | 32.187 ± 0.157 (4552) | 31.8 ± 0.157 (2600) | 31.725 ± 0.395 (647) | 33.234 ± 0.854 (337) | 32.956 ± 0.736 (375) | 31.742 ± 0.735 (207) | 34.148 ± 0.803 (386) |
| Arterial Blood Gas | Base Excess Art | -2.625 ± 0.206 (813) | -2.516 ± 0.436 (131) | -0.594 ± 0.501 (110) | -2.305 ± 0.64 (74) | -2.191 ± 0.426 (153) | -1.205 ± 0.375 (122) | -4.873 ± 0.478 (223) |
| Arterial Blood Gas | CO Hb Art | 0.486 ± 0.021 (334) | 0.52 ± 0.075 (46) | 0.353 ± 0.049 (31) | 0.437 ± 0.055 (29) | 0.432 ± 0.035 (66) | 0.537 ± 0.055 (61) | 0.53 ± 0.04 (101) |
| Arterial Blood Gas | HCO3 Art | 22.284 ± 0.198 (813) | 22.128 ± 0.472 (131) | 24.045 ± 0.585 (110) | 21.985 ± 0.675 (74) | 22.582 ± 0.423 (153) | 23.631 ± 0.381 (122) | 20.665 ± 0.403 (223) |
| Arterial Blood Gas | Hb Art | 12.847 ± 0.13 (334) | 13.259 ± 0.316 (46) | 12.589 ± 0.48 (31) | 13.04 ± 0.443 (29) | 12.919 ± 0.263 (66) | 13.134 ± 0.289 (61) | 12.463 ± 0.259 (101) |
| Arterial Blood Gas | Hct Art (Calc) | 37.795 ± 0.384 (334) | 39.047 ± 0.934 (46) | 36.994 ± 1.408 (31) | 38.339 ± 1.31 (29) | 37.936 ± 0.777 (66) | 38.658 ± 0.852 (61) | 36.702 ± 0.76 (101) |
| Arterial Blood Gas | Ionized Calcium | 1.14 ± 0.001 (2906) | 1.146 ± 0.002 (1572) | 1.141 ± 0.005 (359) | 1.14 ± 0.005 (212) | 1.132 ± 0.006 (286) | 1.115 ± 0.005 (176) | 1.129 ± 0.006 (301) |
| Arterial Blood Gas | Met Hb Art | 0.264 ± 0.009 (334) | 0.264 ± 0.039 (46) | 0.27 ± 0.026 (31) | 0.267 ± 0.027 (29) | 0.256 ± 0.014 (66) | 0.269 ± 0.02 (61) | 0.264 ± 0.015 (101) |
| Arterial Blood Gas | O2 Sat Art | 92.87 ± 0.341 (813) | 92.054 ± 1.431 (131) | 94.466 ± 0.783 (110) | 93.814 ± 1.261 (74) | 93.452 ± 0.581 (153) | 92.697 ± 0.479 (122) | 91.944 ± 0.529 (223) |
| Arterial Blood Gas | O2 Sat Art measured | 92.019 ± 0.485 (334) | 93.047 ± 2.194 (46) | 95.953 ± 0.616 (31) | 95.556 ± 0.733 (29) | 92.363 ± 0.724 (66) | 90.336 ± 1.003 (61) | 90.118 ± 0.892 (101) |
| Arterial Blood Gas | RT Lactate | 1.955 ± 0.018 (5088) | 1.723 ± 0.013 (2951) | 1.901 ± 0.028 (653) | 1.997 ± 0.05 (356) | 2.191 ± 0.065 (426) | 2.147 ± 0.065 (247) | 3.177 ± 0.144 (455) |
| Arterial Blood Gas | pCO2 Art | 39.318 ± 0.41 (813) | 38.464 ± 1.106 (131) | 40.138 ± 1.238 (110) | 36.518 ± 1.275 (74) | 38.983 ± 0.861 (153) | 40.858 ± 0.953 (122) | 39.731 ± 0.796 (223) |
| Arterial Blood Gas | pH Art | 7.372 ± 0.004 (814) | 7.382 ± 0.007 (132) | 7.4 ± 0.007 (110) | 7.398 ± 0.009 (74) | 7.382 ± 0.007 (153) | 7.388 ± 0.007 (122) | 7.329 ± 0.009 (223) |
| Arterial Blood Gas | pO2 Art | 104.623 ± 2.566 (813) | 119.286 ± 8.044 (131) | 113.803 ± 7.001 (110) | 125.866 ± 11.262 (74) | 95.379 ± 4.52 (153) | 89.66 ± 5.348 (122) | 98.962 ± 4.395 (223) |
| Auto Diff | Abs Basophil | 0.022 ± 0 (11134) | 0.024 ± 0 (8080) | 0.018 ± 0.001 (1087) | 0.021 ± 0.001 (573) | 0.02 ± 0.001 (580) | 0.014 ± 0.001 (302) | 0.018 ± 0.001 (512) |
| Auto Diff | Abs Eos | 0.062 ± 0.001 (11134) | 0.071 ± 0.002 (8080) | 0.042 ± 0.003 (1087) | 0.047 ± 0.004 (573) | 0.038 ± 0.004 (580) | 0.026 ± 0.005 (302) | 0.027 ± 0.005 (512) |
| Auto Diff | Abs Immature Grans | 0.061 ± 0.001 (11134) | 0.049 ± 0.001 (8080) | 0.076 ± 0.003 (1087) | 0.077 ± 0.005 (573) | 0.107 ± 0.007 (580) | 0.095 ± 0.008 (302) | 0.115 ± 0.008 (512) |
| Auto Diff | Abs Lymph | 1.303 ± 0.008 (11134) | 1.387 ± 0.009 (8080) | 1.113 ± 0.019 (1087) | 1.204 ± 0.062 (573) | 1.05 ± 0.029 (580) | 0.938 ± 0.036 (302) | 0.989 ± 0.045 (512) |
| Auto Diff | Abs Monocyte | 0.587 ± 0.009 (11134) | 0.596 ± 0.004 (8080) | 0.534 ± 0.009 (1087) | 0.573 ± 0.015 (573) | 0.545 ± 0.014 (580) | 0.485 ± 0.018 (302) | 0.692 ± 0.172 (512) |
| Auto Diff | Abs Neutro | 5.702 ± 0.032 (11134) | 5.336 ± 0.034 (8080) | 6.238 ± 0.106 (1087) | 6.257 ± 0.146 (573) | 6.986 ± 0.171 (580) | 7 ± 0.21 (302) | 7.499 ± 0.196 (512) |
| Auto Diff | Basophil Auto | 0.317 ± 0.002 (11134) | 0.343 ± 0.003 (8080) | 0.26 ± 0.007 (1087) | 0.283 ± 0.01 (573) | 0.254 ± 0.008 (580) | 0.2 ± 0.009 (302) | 0.212 ± 0.008 (512) |
| Auto Diff | Eos Auto | 0.852 ± 0.015 (11134) | 0.99 ± 0.019 (8080) | 0.562 ± 0.038 (1087) | 0.625 ± 0.052 (573) | 0.478 ± 0.053 (580) | 0.302 ± 0.058 (302) | 0.283 ± 0.043 (512) |
| Auto Diff | Immature Grans Auto | 0.705 ± 0.007 (11134) | 0.615 ± 0.008 (8080) | 0.868 ± 0.026 (1087) | 0.855 ± 0.037 (573) | 1.048 ± 0.048 (580) | 0.963 ± 0.059 (302) | 1.074 ± 0.049 (512) |
| Auto Diff | Lymph Auto | 18.966 ± 0.1 (11134) | 20.6 ± 0.118 (8080) | 15.871 ± 0.278 (1087) | 16.017 ± 0.391 (573) | 13.913 ± 0.347 (580) | 12.716 ± 0.489 (302) | 12.459 ± 0.407 (512) |
| Auto Diff | Mono Auto | 8.076 ± 0.037 (11134) | 8.529 ± 0.043 (8080) | 7.034 ± 0.102 (1087) | 7.516 ± 0.166 (573) | 6.682 ± 0.137 (580) | 6.034 ± 0.189 (302) | 6.535 ± 0.234 (512) |
| Auto Diff | Neutro Auto | 71.076 ± 0.119 (11134) | 68.91 ± 0.138 (8080) | 75.405 ± 0.336 (1087) | 74.704 ± 0.475 (573) | 77.626 ± 0.428 (580) | 79.787 ± 0.587 (302) | 79.438 ± 0.525 (512) |
| BNP | B-Type Natri Peptide | 269.762 ± 13.803 (3343) | 209.708 ± 14.23 (1785) | 284.654 ± 39.548 (479) | 335.649 ± 56.311 (255) | 274.791 ± 34.34 (298) | 398.893 ± 115.72 (164) | 437.126 ± 57.851 (362) |
| C Reactive Protein | C Reactive Prot | 10.04 ± 0.117 (5406) | 7.755 ± 0.136 (2968) | 11.327 ± 0.283 (835) | 11.567 ± 0.451 (413) | 13.62 ± 0.435 (453) | 14.738 ± 0.557 (271) | 14.722 ± 0.438 (466) |
| CBC with Diff | Automated Nucleated RBC's | 0.059 ± 0.011 (12557) | 0.044 ± 0.013 (8924) | 0.093 ± 0.052 (1277) | 0.024 ± 0.005 (655) | 0.098 ± 0.025 (683) | 0.084 ± 0.017 (368) | 0.174 ± 0.034 (650) |
| CBC with Diff | Hct | 39.46 ± 0.051 (12654) | 39.836 ± 0.057 (9014) | 38.959 ± 0.171 (1279) | 38.507 ± 0.242 (658) | 38.096 ± 0.252 (685) | 39.528 ± 0.333 (368) | 37.598 ± 0.271 (650) |
| CBC with Diff | Hgb | 12.959 ± 0.019 (12654) | 13.119 ± 0.021 (9014) | 12.752 ± 0.061 (1279) | 12.568 ± 0.087 (658) | 12.423 ± 0.09 (685) | 12.918 ± 0.115 (368) | 12.119 ± 0.093 (650) |
| CBC with Diff | MCH | 29.309 ± 0.023 (12653) | 29.284 ± 0.027 (9013) | 29.359 ± 0.073 (1279) | 29.269 ± 0.093 (658) | 29.321 ± 0.105 (685) | 29.325 ± 0.125 (368) | 29.577 ± 0.099 (650) |
| CBC with Diff | MCHC | 32.777 ± 0.013 (12647) | 32.875 ± 0.015 (9008) | 32.666 ± 0.043 (1279) | 32.561 ± 0.059 (657) | 32.515 ± 0.061 (685) | 32.654 ± 0.079 (368) | 32.199 ± 0.062 (650) |
| CBC with Diff | MCV | 89.382 ± 0.056 (12654) | 89.029 ± 0.064 (9014) | 89.826 ± 0.183 (1279) | 89.867 ± 0.238 (658) | 90.081 ± 0.262 (685) | 89.817 ± 0.334 (368) | 91.932 ± 0.276 (650) |
| CBC with Diff | MPV | 10.482 ± 0.009 (12540) | 10.448 ± 0.01 (8941) | 10.483 ± 0.026 (1267) | 10.523 ± 0.039 (649) | 10.55 ± 0.037 (681) | 10.581 ± 0.048 (361) | 10.779 ± 0.04 (641) |
| CBC with Diff | Platelet | 231.18 ± 0.819 (12654) | 233.928 ± 0.937 (9014) | 231.689 ± 2.755 (1279) | 229.468 ± 4.032 (658) | 229.112 ± 4.031 (685) | 218.107 ± 4.647 (368) | 203.38 ± 3.607 (650) |
| CBC with Diff | RBC | 4.435 ± 0.006 (12654) | 4.492 ± 0.007 (9014) | 4.359 ± 0.021 (1279) | 4.304 ± 0.029 (658) | 4.256 ± 0.031 (685) | 4.425 ± 0.041 (368) | 4.119 ± 0.032 (650) |
| CBC with Diff | RDW | 13.784 ± 0.017 (12647) | 13.612 ± 0.02 (9008) | 13.987 ± 0.058 (1279) | 14.109 ± 0.078 (658) | 14.346 ± 0.085 (685) | 14.087 ± 0.099 (368) | 14.672 ± 0.085 (649) |
| CBC with Diff | WBC | 7.888 ± 0.073 (12654) | 7.533 ± 0.085 (9014) | 8.003 ± 0.155 (1279) | 8.294 ± 0.178 (658) | 8.905 ± 0.19 (685) | 8.952 ± 0.273 (368) | 10.512 ± 0.662 (650) |
| Comprehensive Metabolic Panel | AGAP | 11.298 ± 0.025 (12355) | 11.065 ± 0.027 (8702) | 11.359 ± 0.073 (1281) | 11.631 ± 0.112 (653) | 11.812 ± 0.115 (688) | 11.787 ± 0.152 (369) | 13.111 ± 0.193 (662) |
| Comprehensive Metabolic Panel | ALT | 46.505 ± 1.222 (11370) | 40.93 ± 0.656 (7915) | 51.823 ± 4.232 (1210) | 45.727 ± 3.123 (618) | 54.862 ± 8.657 (646) | 50.59 ± 5.623 (350) | 96.176 ± 15.791 (631) |
| Comprehensive Metabolic Panel | AST | 53.579 ± 2.147 (11512) | 41.915 ± 0.641 (8018) | 58.88 ± 4.4 (1223) | 56.349 ± 4.847 (628) | 79.851 ± 16.943 (653) | 61.92 ± 3.621 (353) | 155.926 ± 31.982 (637) |
| Comprehensive Metabolic Panel | Albumin Level | 3.339 ± 0.006 (11454) | 3.502 ± 0.007 (7965) | 3.073 ± 0.016 (1219) | 3.063 ± 0.025 (627) | 2.926 ± 0.022 (653) | 2.872 ± 0.028 (353) | 2.762 ± 0.023 (637) |
| Comprehensive Metabolic Panel | Alk Phos | 91.93 ± 0.56 (11448) | 89.848 ± 0.633 (7960) | 94.316 ± 1.792 (1219) | 98.619 ± 2.94 (627) | 96.766 ± 2.544 (652) | 96.011 ± 3.661 (353) | 99.59 ± 2.708 (637) |
| Comprehensive Metabolic Panel | BUN | 17.698 ± 0.141 (12357) | 15.003 ± 0.127 (8704) | 19.877 ± 0.483 (1281) | 22.556 ± 0.779 (654) | 23.975 ± 0.801 (687) | 23.197 ± 0.934 (369) | 34.54 ± 0.989 (662) |
| Comprehensive Metabolic Panel | Bili Total | 0.643 ± 0.007 (11436) | 0.609 ± 0.007 (7950) | 0.697 ± 0.026 (1219) | 0.694 ± 0.043 (626) | 0.697 ± 0.029 (652) | 0.66 ± 0.032 (353) | 0.847 ± 0.053 (636) |
| Comprehensive Metabolic Panel | CO2 | 22.731 ± 0.032 (12355) | 22.853 ± 0.035 (8702) | 22.878 ± 0.107 (1281) | 22.428 ± 0.152 (653) | 22.486 ± 0.155 (688) | 22.478 ± 0.196 (369) | 21.542 ± 0.192 (662) |
| Comprehensive Metabolic Panel | Calcium | 8.854 ± 0.006 (12356) | 8.944 ± 0.006 (8703) | 8.712 ± 0.017 (1281) | 8.713 ± 0.026 (653) | 8.627 ± 0.027 (688) | 8.498 ± 0.033 (369) | 8.519 ± 0.029 (662) |
| Comprehensive Metabolic Panel | Chloride | 102.797 ± 0.041 (12356) | 103.039 ± 0.045 (8703) | 102.101 ± 0.137 (1281) | 102.348 ± 0.202 (653) | 102.063 ± 0.217 (688) | 101.687 ± 0.249 (369) | 102.778 ± 0.256 (662) |
| Comprehensive Metabolic Panel | Creatinine | 1.222 ± 0.013 (12429) | 1.11 ± 0.013 (8771) | 1.303 ± 0.044 (1284) | 1.516 ± 0.076 (655) | 1.437 ± 0.065 (688) | 1.464 ± 0.092 (369) | 1.899 ± 0.08 (662) |
| Comprehensive Metabolic Panel | Glucose Level | 141.359 ± 0.66 (12371) | 132.074 ± 0.715 (8717) | 154.543 ± 2.149 (1281) | 156.805 ± 3.17 (654) | 165.604 ± 3.219 (688) | 177.692 ± 4.07 (369) | 177.405 ± 3.694 (662) |
| Comprehensive Metabolic Panel | Potassium Level | 3.952 ± 0.004 (12362) | 3.904 ± 0.005 (8709) | 4.012 ± 0.014 (1281) | 4.017 ± 0.021 (653) | 4.05 ± 0.02 (687) | 4.109 ± 0.025 (369) | 4.199 ± 0.024 (663) |
| Comprehensive Metabolic Panel | Sodium Level | 136.832 ± 0.034 (12358) | 136.962 ± 0.037 (8704) | 136.342 ± 0.113 (1282) | 136.404 ± 0.164 (653) | 136.377 ± 0.189 (688) | 135.971 ± 0.213 (369) | 137.443 ± 0.232 (662) |
| Comprehensive Metabolic Panel | Total Protein | 7.254 ± 0.007 (11449) | 7.349 ± 0.008 (7961) | 7.125 ± 0.02 (1218) | 7.136 ± 0.03 (627) | 7.045 ± 0.031 (652) | 7.055 ± 0.04 (353) | 6.755 ± 0.035 (638) |
| D-Dimer | D-Dimer | 2153.8 ± 81.72 (6271) | 1462.169 ± 67.832 (3743) | 2427.967 ± 243.774 (886) | 2448.171 ± 296.275 (417) | 3068.07 ± 385.886 (449) | 4028.402 ± 665.225 (278) | 4747.1 ± 515.16 (498) |
| Fibrinogen Level | Fibrinogen | 510.385 ± 4.139 (1726) | 486.914 ± 5.226 (951) | 537.043 ± 10.22 (257) | 517.167 ± 15.489 (132) | 548.382 ± 16.824 (126) | 566.292 ± 18.179 (110) | 534.638 ± 15.781 (150) |
| Glomerular Filtration Rate | GFR (AA) | 38.556 ± 0.353 (2306) | 40.161 ± 0.492 (1197) | 37.533 ± 0.972 (291) | 36.523 ± 1.29 (186) | 37.633 ± 1.177 (204) | 38.06 ± 1.909 (86) | 35.586 ± 0.867 (342) |
| Glomerular Filtration Rate | GFR (NAA) | 40.068 ± 0.279 (3403) | 42.18 ± 0.359 (1924) | 38.398 ± 0.816 (404) | 37.916 ± 1.068 (256) | 37.869 ± 0.996 (271) | 40.362 ± 1.487 (130) | 34.614 ± 0.778 (418) |
| Prothrombin Time with INR | INR | 1.231 ± 0.006 (5330) | 1.197 ± 0.008 (3074) | 1.237 ± 0.015 (754) | 1.273 ± 0.024 (390) | 1.295 ± 0.026 (432) | 1.228 ± 0.031 (229) | 1.355 ± 0.029 (451) |
| Prothrombin Time with INR | PT | 14.163 ± 0.07 (5330) | 13.8 ± 0.087 (3074) | 14.225 ± 0.158 (754) | 14.62 ± 0.267 (390) | 14.853 ± 0.287 (432) | 14.103 ± 0.329 (229) | 15.513 ± 0.313 (451) |
| Sedimentation Rate | Sed Rate | 42.278 ± 1.498 (465) | 35.633 ± 1.822 (282) | 54.188 ± 3.231 (85) | 51.556 ± 6.163 (36) | 55.861 ± 5.535 (36) | 46.5 ± 11.637 (10) | 42.062 ± 7.544 (16) |
| Troponin I | Troponin-I | 0.315 ± 0.069 (4824) | 0.196 ± 0.06 (2827) | 0.114 ± 0.049 (610) | 0.44 ± 0.242 (330) | 0.264 ± 0.06 (375) | 1.038 ± 0.87 (215) | 0.917 ± 0.419 (467) |
| Venous Blood Gas | Base Excess Ven | -1.021 ± 0.066 (4480) | -0.733 ± 0.073 (2623) | -0.523 ± 0.178 (591) | -1.132 ± 0.256 (300) | -1.259 ± 0.258 (386) | -1.182 ± 0.303 (205) | -3.399 ± 0.359 (375) |
| Venous Blood Gas | HCO3 Ven | 23.951 ± 0.065 (4480) | 24.147 ± 0.075 (2623) | 24.416 ± 0.189 (591) | 23.785 ± 0.267 (300) | 23.74 ± 0.251 (386) | 23.639 ± 0.311 (205) | 22.365 ± 0.285 (375) |
| Venous Blood Gas | O2 Sat Ven | 58.895 ± 0.327 (4477) | 59.187 ± 0.425 (2621) | 58.662 ± 0.888 (590) | 60.945 ± 1.267 (300) | 58.804 ± 1.075 (386) | 58.544 ± 1.531 (205) | 55.87 ± 1.204 (375) |
| Venous Blood Gas | RT Chloride | 100.665 ± 0.105 (2571) | 100.693 ± 0.122 (1382) | 100.404 ± 0.311 (326) | 100.066 ± 0.41 (185) | 100.205 ± 0.427 (249) | 100.214 ± 0.37 (161) | 101.954 ± 0.425 (268) |
| Venous Blood Gas | RT Glucose | 166.425 ± 1.927 (2593) | 154.511 ± 2.528 (1396) | 162.996 ± 5.236 (328) | 182.747 ± 7.582 (186) | 187.09 ± 6.873 (251) | 186.925 ± 7.018 (164) | 189.455 ± 6.199 (268) |
| Venous Blood Gas | RT Potassium | 4.023 ± 0.013 (2882) | 3.948 ± 0.016 (1565) | 4.073 ± 0.039 (356) | 4.06 ± 0.046 (209) | 4.061 ± 0.046 (278) | 4.06 ± 0.055 (175) | 4.271 ± 0.048 (299) |
| Venous Blood Gas | RT Sodium | 135.327 ± 0.097 (2876) | 135.433 ± 0.115 (1564) | 135 ± 0.276 (357) | 134.524 ± 0.309 (209) | 134.845 ± 0.395 (277) | 134.64 ± 0.357 (173) | 136.579 ± 0.424 (296) |
| Venous Blood Gas | pCO2 Ven | 40.971 ± 0.128 (4480) | 40.77 ± 0.143 (2623) | 41.356 ± 0.389 (591) | 40.557 ± 0.487 (300) | 40.778 ± 0.471 (386) | 40.095 ± 0.572 (205) | 42.772 ± 0.685 (375) |
| Venous Blood Gas | pH Ven | 7.385 ± 0.001 (4485) | 7.391 ± 0.001 (2626) | 7.39 ± 0.003 (591) | 7.385 ± 0.004 (301) | 7.381 ± 0.004 (386) | 7.388 ± 0.004 (205) | 7.335 ± 0.008 (376) |
| Venous Blood Gas | pO2 Ven | 35.46 ± 0.292 (4475) | 34.407 ± 0.264 (2619) | 34.259 ± 0.561 (589) | 36.538 ± 0.987 (300) | 36.533 ± 1.24 (386) | 43.18 ± 3.09 (205) | 38.505 ± 1.641 (376) |

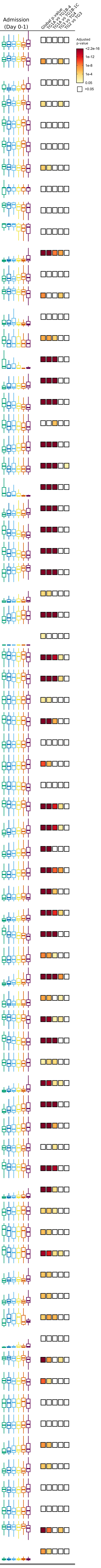

### Table S9 - Vital Signs by Trajectory Group

| Vital Sign | Total (21312) | Moderate | Severe | | Critical | | Death |
|---|---|---|---|---|---|---|---|
| | | TG1A (17476) | TG1B (1351) | TG1C (701) | TG2 (715) | TG3 (380) | TG4 (689) |
| Glasgow Coma Score | 14.61 ± 0.015 (11380) | 14.894 ± 0.007 (8565) | 14.568 ± 0.046 (920) | 14.393 ± 0.07 (502) | 13.725 ± 0.113 (551) | 12.748 ± 0.222 (308) | 12.316 ± 0.174 (534) |
| Oxygen Saturation | 96.076 ± 0.021 (17319) | 96.719 ± 0.019 (13512) | 94.156 ± 0.073 (1345) | 94.623 ± 0.109 (690) | 93.964 ± 0.107 (712) | 92.706 ± 0.167 (379) | 92.662 ± 0.146 (681) |
| Peripheral Pulse Rate | 87.655 ± 0.124 (15895) | 87.124 ± 0.135 (12614) | 88.333 ± 0.45 (1217) | 89.381 ± 0.684 (606) | 91.337 ± 0.714 (621) | 92.13 ± 1.025 (302) | 89.862 ± 0.839 (535) |
| Respiratory Rate | 19.172 ± 0.029 (17312) | 18.407 ± 0.025 (13506) | 20.828 ± 0.117 (1347) | 21.13 ± 0.19 (692) | 22.061 ± 0.2 (708) | 24.032 ± 0.296 (377) | 23.379 ± 0.223 (682) |
| Temperature Oral (DegC) | 37.118 ± 0.004 (16198) | 37.126 ± 0.004 (12730) | 37.11 ± 0.015 (1290) | 37.09 ± 0.022 (657) | 37.091 ± 0.022 (642) | 37.197 ± 0.03 (331) | 36.985 ± 0.022 (548) |
| Diastolic Blood Pressure | 76.141 ± 0.079 (17414) | 77.005 ± 0.09 (13624) | 74.49 ± 0.255 (1346) | 74.313 ± 0.376 (692) | 71.991 ± 0.342 (711) | 71.974 ± 0.458 (369) | 70.477 ± 0.378 (672) |
| Systolic Blood Pressure | 128.692 ± 0.136 (17414) | 128.968 ± 0.153 (13624) | 128.569 ± 0.486 (1346) | 129.943 ± 0.721 (692) | 126.813 ± 0.655 (711) | 125.479 ± 0.903 (369) | 125.805 ± 0.773 (672) |

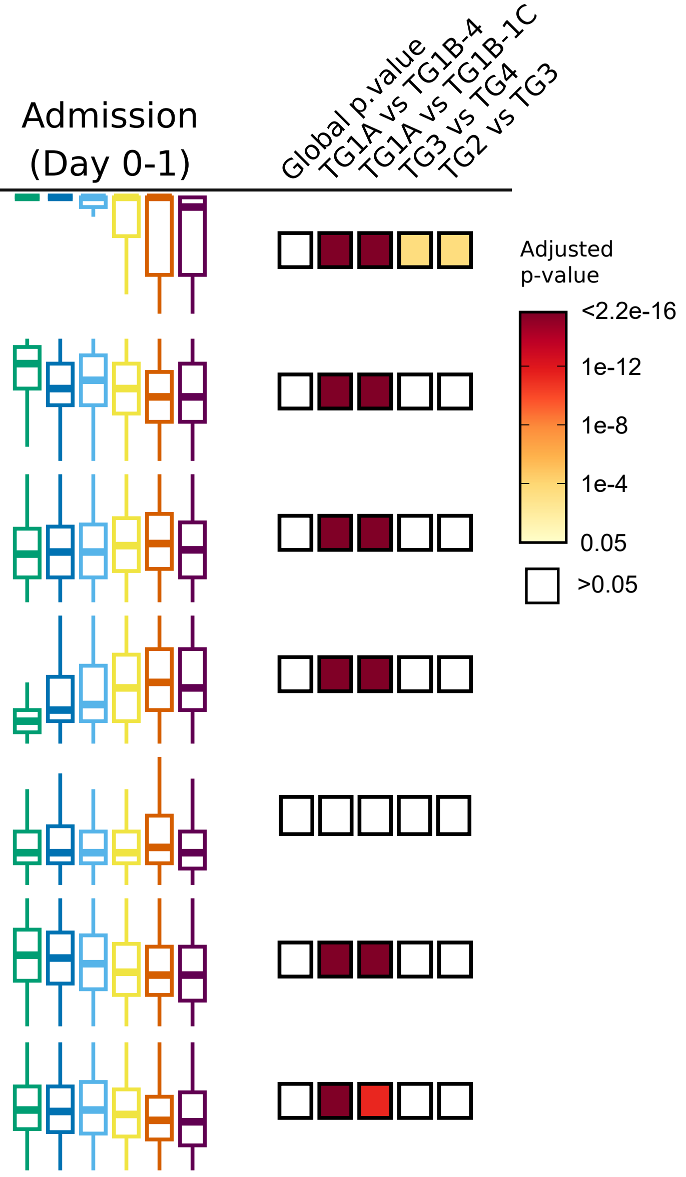
